# Course of Itch from Systemic Sclerosis Onset: a Scleroderma Patient-Centred Intervention Network Cohort Longitudinal Study

**DOI:** 10.64898/2026.03.31.26349869

**Authors:** Meira Golberg, Marie-Eve Carrier, Gil Yosipovitch, Cassidy Dal Santo, Linda Kwakkenbos, Tracy Frech, Sabrina Hoa, Elena Netchiporouk, Laurent Misery, Jo-Ann Lapointe McKenzie, Tracy Mieszczak, Sandra Rideout, Maureen Sauvé, Anie Philip, Janet Pope, Susan J. Bartlett, Benjamin Chaigne, Catherine Fortuné, Amy Gietzen, Karen Gottesman, Geneviève Guillot, Laura K. Hummers, Amanda Lawrie-Jones, Vanessa L. Malcarne, Maureen D. Mayes, Yanne Perriault, Danielle Rice, Michelle Richard, James Stempel, Robyn K. Wojeck, Luc Mouthon, Andrea Benedetti, Brett D. Thombs, the Scleroderma Patient-centered Intervention Network Investigators

**Affiliations:** Lady Davis Institute for Medical Research, Jewish General Hospital, Montréal, Québec, Canada; Department of Dermatology and Cutaneous Surgery, University of Miami Miller School of Medicine, Coral Gables, Florida, United States; Department of Educational and Counselling Psychology, McGill University, Montréal, Québec, Canada; Department of Clinical Psychology, Behavioural Science Institute, Radboud University, Nijmegen, the Netherlands; Centre for Mindfulness, Department of Psychiatry, Radboud University Medical Center, Nijmegen, the Netherlands; Division of Rheumatology and Immunology, Vanderbilt University Medical Center, Nashville, Tennessee, United States; Tennessee Valley Health Care System, Veterans Affair Medical Center, Nashville, Tennessee, United States; Centre Hospitalier de l’Université de Montréal, Montréal, Québec, Canada; Département de Médecine, Division de Rhumatologie, Université de Montréal, Montréal, Québec, Canada; Division of Dermatology, Department of Medicine, McGill University Health Centre, Montréal, Québec, Canada; Research Institute of the McGill University Health Centre, Montréal, Québec, Canada; Dermatology Department, Brest University Hospital, Brest, France; Scleroderma Manitoba, Oak Bluff, Manitoba, Canada; Scleroderma Patient-centered Intervention Network, Rochester, New York, United States; Scleroderma Atlantic, Halifax, Nova Scotia, Canada; Scleroderma Society of Ontario, Hamilton, Ontario, Canada; Scleroderma Canada, Hamilton, Ontario, Canada; Department of Surgery, McGill University, Montréal, Québec, Canada; SkinInvestigation Network of Canada, Toronto, Ontario, Canada; Department of Medicine, Division of Dermatology, Western University, London, Ontario, Canada; Department of Medicine, McGill University, Montréal, Québec, Canada; Service de Médecine Interne, Centre de Référence Maladies Autoimmunes Systémiques Rares d’Ile de France, Hôpital Cochin, Paris, France; Assistance Publique Hôpitaux de Paris-Centre, Hôpital Cochin, Université Paris Cité, Paris, France; Ottawa Scleroderma Support Group, Ottawa, Ontario, Canada; Steffens Scleroderma Foundation, Albany, New York, United States; National Scleroderma Foundation, Los Angeles, California, United States; Sclérodermie Québec, Longueuil, Québec, Canada; Johns Hopkins University School of Medicine, Baltimore, Maryland, United States; Scleroderma Australia, Melbourne, Victoria, Australia; Scleroderma Victoria, Melbourne, Victoria, Australia; Department of Psychology, San Diego State University, San Diego, California, United States; San Diego State University/University of California, San Diego Joint Doctoral Program in Clinical Psychology, San Diego, California, United States; University of Texas McGovern School of Medicine, Houston, Texas, United States; Association des sclérodermiques de France, Baccon, France; Department of Psychology, St. Joseph’s Healthcare Hamilton, Hamilton, Ontario, Canada; Department of Psychiatry & Behavioural Neurosciences, McMaster University, Hamilton, Ontario, Canada; Scleroderma Foundation of Greater Chicago, Chicago, Illinois, United States; Amgen Inc, Thousand Oaks, California, United States; Department of Epidemiology, Biostatistics, and Occupational Health, McGill University, Montréal, Québec, Canada; Respiratory Epidemiology and Clinical Research Unit, McGill University Health Centre, Montréal, Québec, Canada; Department of Psychiatry, McGill University, Montréal, Québec, Canada

**Author notes:** Corresponding author: Brett D. Thombs, PhD; Jewish General Hospital; 3755 Côte Ste Catherine Road, Pavilion H4.83, Montréal, Québec, Canada, H3T 1E2.

## Abstract

**Background:** Itch in systemic sclerosis (SSc) is thought to be most significant in early disease, but no longitudinal studies have examined itch course. We estimated itch presence and severity from SSc disease onset, accounting for participant age and time since onset at each assessment.

**Methods:** People with SSc from the multinational Scleroderma Patient-centred Intervention Network Cohort completed past-week itch severity assessments (0 to 10 numerical rating scale) at enrolment and longitudinally at 3-month intervals. To estimate itch probability (score > 0) and, if present, itch severity, we used two-stage mixed effects models with basis splines to address non-linearity. The primary predictor was age at each assessment, partitioned into age at non-Raynaud phenomenon symptom onset and time since onset. We estimated prevalence and severity for onset ages of 20, 30, 40, 50 and 60 years and, for each onset age, at 2 years, 3 years, 4 years, 5 years, 7 years, and 5-year intervals 10 years to 35 years post-onset.

**Findings:** We included 2173 participants with 19 733 itch assessments (mean [standard deviation] 9·1 [6·9] assessments). 1896 of 2173 (87·3%) participants were women. Mean age at enrolment was 54·7 (SD 12·7) years. 873 (40·2%) participants had diffuse cutaneous SSc. Predicted itch probability was between 35·0% (95% CI 31·8% to 38·5%) and 36·8% (95% CI 33·3% to 40·4%) at all onset age and disease duration combinations. Mean itch severity, when present, was moderate, between 4·1 (95% CI 4·1 to 4·1) and 4·4 (95% CI 4·3 to 4·4), for all age and duration combinations.

**Interpretation:** Itch prevalence and mean severity were stable across onset ages and over time within onset ages. Findings suggest that itch is common in SSc and not as closely related to disease duration as previously thought. Research is needed to elucidate itch pathophysiology and identify effective management strategies.

**Funding:** Funding for the study was provided by a Skin Investigation Network of Canada Team Development Award. Funding for the Scleroderma Patient-centred Intervention Network Cohort has been received from the Canadian Institutes of Health Research (TR3-119192; PJT-149073; PJT-148504; PJT-195879; PJT-203755); the Arthritis Society; the Lady Davis Institute for Medical Research of the Jewish General Hospital, Montréal, Québec, Canada; the Jewish General Hospital Foundation, Montréal, Québec, Canada; McGill University, Montréal, Québec, Canada Scleroderma Society of Ontario; Scleroderma Canada; Sclérodermie Québec; Scleroderma Manitoba; Scleroderma Atlantic; the Scleroderma Association of BC; Scleroderma SASK; Scleroderma Australia; Scleroderma New South Wales; Scleroderma Victoria; and the Scleroderma Foundation of California.

**RESEARCH IN CONTEXT:** *Evidence before this study:* We searched PubMed using the terms “itch” or “pruritus” with “systemic sclerosis” or “scleroderma” on March 26, 2025, to identify previous studies that have evaluated the trajectory of itch prevalence or severity in systemic sclerosis (SSc) from the time of disease onset. We did not find any longitudinal studies. We identified 4 cross-sectional studies, and none found statistically significant associations between disease duration and itch. Three of the studies included between 56 and 126 participants. The fourth study included 959 participants and found that itch was experienced on most days in the last month based on a single dichotomous item among 46% of participants between 1 and 4·9 years since non-Raynaud phenomenon (non-RP) symptom onset and 41% for those 5 or more years since onset (not statistically significant).

*Added value of this study:* This was the first longitudinal study of itch prevalence and severity in SSc. We evaluated 2173 Scleroderma Patient-Centred Intervention Network participants from 7 countries who reported itch severity in the past week (0 to 10 numerical rating scale) at cohort enrolment and subsequently at 3-month intervals (19 733 total itch assessments). We simultaneously modelled probability of having any itch and, if present, itch severity. We accounted for both normal aging and SSc disease duration by including age of onset of non-RP symptoms and time since onset in our models. We found that itch prevalence and mean severity were stable across the course of the disease. Between 35% and 37% of participants reported itch (numerical rating scale score > 0) across all ages of onset and time since onset combinations. Mean itch severity, among participants with itch, was between 4·1 and 4·4 points, a moderate level, at all onset age and disease duration combinations. Findings were consistent for subgroups defined by participant country, sex, and diffuse versus limited cutaneous SSc.

*Implications of all the available evidence:* Itch is rarely researched in SSc, and itch assessment and management are typically not part of routine SSc care. It is commonly assumed that itch is most prominent, if present, in early disease. Our study showed that, contrary to this assumption, itch is present for many people with SSc across the course of the disease; itch prevalence and mean severity were stable across time regardless of age of SSc onset. Findings from our study underline the need for research on the pathogenesis of itch in SSc and the development and testing of treatments. Itch assessment and management should be part of routine SSc care.

## INTRODUCTION

Systemic sclerosis (scleroderma, SSc) is a rare, complex, autoimmune connective tissue disease that affects the skin and multiple organ systems, including the lungs, heart, kidneys, musculoskeletal system, and gastrointestinal tract.^1^ Common manifestations that impair function and reduce quality of life include hand function and mobility limitations, fatigue, gastrointestinal symptoms, pain, and itch.^2–5^

Two studies with ≥ 100 participants have reported itch prevalence in SSc. One found that 43% of 959 participants recruited from Canadian outpatient rheumatology clinics experienced itch on most days in the past month based on a single item.^3,6^ The other reported that 44% of 126 participants from an Italian outpatient dermatology clinic reported itch at least sometimes in the last week.^7^ Canadian outpatients (N = 578) with itch most days in the last month had substantially worse mental and physical function and greater disability compared to participants without itch on most days.^4^

Itch prevalence increases with age among adults in the general population.^8,9^ In SSc, itch is thought to be most significant in early disease when activity is greatest.^5,10^ No longitudinal studies have evaluated itch course in SSc. Several cross-sectional studies have examined itch prevalence or severity by disease duration, but none were designed to disentangle possible effects of aging and SSc disease duration, and none found a statistically significant association between disease duration and itch.^3,7,11,12^

Our objective was to estimate the trajectory of past-week itch presence and severity from disease onset among people with SSc, accounting for participant age and time since non-Raynaud phenomenon (non-RP) symptom onset at each assessment.

## METHODS

In this multicentre study, we evaluated longitudinal data from participants in the Scleroderma Patient-centred Intervention Network (SPIN) Cohort.^13^ We posted a protocol prior to initiating the study (https://osf.io/6z5wt/files/dpqmz). We report results based on the Strengthening the Reporting of Observational Studies in Epidemiology statement.^14^ Methods from studies that use SPIN Cohort data are similar. Thus, we followed Text Recycling Research Project reporting guidance.^15^

### Participants and Procedures

SPIN Cohort participants are recruited from sites in Australia, Canada, France, Mexico, Spain, the UK, and the USA. They must be aged ≥ 18 years; fluent in English, French, or Spanish; and classified as having SSc based on 2013 American College of Rheumatology/European League Against Rheumatism criteria^16^ by a SPIN site physician. The SPIN Cohort is a convenience sample. Eligible participants are recruited by attending physicians or nurse coordinators during outpatient visits. Site personnel obtain written informed consent and submit an online medical data form. An automated email is sent to participants with instructions to activate their online SPIN account and complete study measures. SPIN Cohort participants are invited to complete outcome measures via an online portal upon enrolment and subsequently every three months. The SPIN Cohort study was approved by the Research Ethics Committee of the Centre intégré universitaire de santé et de services sociaux du Centre-Ouest-de-l’Île-de-Montréal (#MP-05-2013-150) and by ethics committees of all recruiting sites. Participant recruitment is ongoing.

The present study included data at the time of enrolment and follow-up assessments from cohort inception in April 2014 to October 10, 2020, when assessment of itch was discontinued. Participants were included if they (1) completed an item about past-week itch at least once and (2) had physician-reported time since non-RP symptom onset at enrolment.

### Measures

#### Sociodemographic and Medical Variables

SPIN Cohort participants provided sociodemographic (race or ethnicity, years of education, marital status) and lifestyle (smoking status, alcohol consumption) information and completed patient-reported outcome measures. Site physicians, at enrolment, provided participant age, sex, height, weight, date of initial onset of non-RP symptoms, SSc subtype (limited cutaneous, diffuse cutaneous, sine scleroderma), modified Rodnan skin score,^17^ presence of gastrointestinal symptoms (upper, lower, none), presence of digital ulcers anywhere on the fingers, presence of tendon friction rubs (currently, in the past, never), presence of small or large joint contractures (none, mild [≤ 25% range-of-motion limitation], moderate to severe [> 25% range-of-motion limitation]), presence of pulmonary arterial hypertension, presence of interstitial lung disease, history of scleroderma renal crisis, presence of current or past overlap syndromes or associations (systemic lupus erythematosus, rheumatoid arthritis, Sjögren’s disease, autoimmune thyroid disease, idiopathic inflammatory myopathy, primary biliary cholangitis), and presence of SSc-related antibodies (antinuclear antibody with nucleolar pattern, anticentromere, antitopoisomerase I, anti-RNA polymerase III).

#### Itch Numerical Rating Scale

Past-week itch severity was measured with a single-item numerical rating scale (*How severe was your itch in the past week?*; 0 = not severe at all to 10 = unbearable). Multiple single-item numerical rating scales, scored 0 to 10, have been used to assess itch and, despite item wording differences, consistently generate good test-retest reliability (intraclass correlation coefficient > 0·70)^18,19^ and demonstrate convergent validity with multi-item itch scales (Pearson’s correlations 0·61 to 0·77).^18^ They correlate well (Spearman’s correlation > 0·80) with other validated single-item itch severity measures, including visual analogue and verbal rating scales.^19^ Differences of 3 to 4 points have been used to reflect a minimally important difference.^20^

### Statistical Analysis

We described participant characteristics at SPIN Cohort enrolment using frequencies and percentages for categorical variables and means and standard deviations for continuous variables. To estimate the probability of experiencing any itch (numerical rating scale score > 0) and, separately, if present, itch severity, we used two-stage mixed effects models. The two-stage approach addressed the high proportion of zero scores by separately modelling presence of itch via a generalized linear mixed effects model with a logistic link and itch severity as a linear mixed effects model.^21–23^ The primary predictor in each model was age at time of assessment, which was partitioned into age at onset of non-RP symptoms and time since onset of non-RP symptoms. In each model, we estimated itch status *Y* for individual *i* at assessment time *t*.

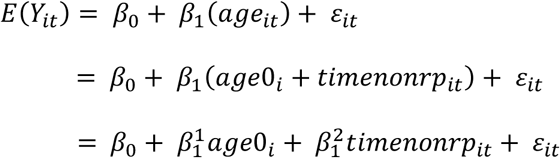

where *age_it_* represented age for individual *i* at time of assessment *t*, *age0_i_* represented age of non-RP symptom onset for individual *i*, and *timenonrp_it_* represented time since non-RP symptom onset for individual *i* at time of assessment *t*.

We accounted for non-RP symptom onset age and time since non-RP symptom onset separately because age at time of assessment increases across assessments at the same rate as time since non-RP onset. Thus, time-varying variables for both age and time since non-RP symptom would be fully collinear. Including a single term for time, without differentiating, on the other hand, would not allow evaluation of whether trajectories differ depending on age of onset or determination of whether within-person changes in itch over time are associated with chronological aging versus increasing time since non-RP symptom onset.^24^ We included random intercepts for participants to account for correlations between repeated measures within persons. We also adjusted for country (Canada, France, UK, other [Australia, Mexico, Spain]; reference = USA), male sex (reference = female), and diffuse cutaneous SSc (reference = limited cutaneous SSc or sine scleroderma). We adjusted for a small set of covariates that are stable over time for most people with SSc and did not include time-varying predictors. Medical variables are collected in the SPIN Cohort only at the time of enrolment, which is not aligned with the time of non-RP symptom onset, and our objective was to map itch trajectory and not to attempt to identify factors that may contribute to itch.

We anticipated that trajectories of itch may not be linear. Thus, we incorporated basis splines with the R *splines* package (version 4·2·2), which allowed us to flexibly represent non-linear changes in itch.^25,26^ To allow itch to vary nonlinearly both (1) between individuals with different ages of non-RP symptom onset and (2) within individuals over time since non-RP symptom onset, we considered basis splines for both variables. Basis splines consist of piecewise polynomial functions of user-specified degree. By default, knots are placed at the minimum, median, and maximum values. We used the Akaike Information Criterion (AIC) to determine the degree of polynomials and number of knots to retain. We defined knot locations in advance based on the quantiles of age of non-RP symptom onset and time since non-RP symptom onset, considering a maximum of five knots for each.^25,26^ We selected the lowest degree model which did not substantially increase AIC. To ensure we retained sufficiently complex models without overfitting, we also visually inspected fitted model trajectories with varying complexity but similar AIC.

We estimated trajectories of past week probability of itch and, if present, severity for ages at time of assessment from 20 years to 80 years. We also estimated probability of past week itch presence and, if present, severity with 95% confidence intervals (CIs) at two years, three years, four years, five years, seven years, and at 5-year intervals from ten years to 35 years post non-RP symptom onset. To evaluate whether trajectories of itch differed based on age of non-RP symptom onset, we tested the interaction of age at onset of non-RP symptoms and time since onset. We also reported trajectories and predicted values separately for non-RP symptom onset ages of 20 years, 30 years, 40 years, 50 years, and 60 years. For each predicted onset-age trajectory, we defined reported boundaries using all participants with actual onset age ± 5 years from the prediction age (e.g., ≥25 to <35 years for 30-year estimated trajectory). We started each onset age trajectory at the 10^th^ percentile of time since non-RP symptom onset and finished at the 90^th^ percentile for participants in the trajectory to ensure adequate data for estimates. Modelling was done using the GLMMAdaptive package (version 4·2·2).^22^

We conducted a complete data analysis rather than imputing to attempt to account for missing data. This is because imputation would have required the generation of a large proportion of model data when considering longitudinal trajectories between the time of non-RP symptom onset and SPIN Cohort enrolment for many participants.

Separately from the main analyses, we showed results from all analyses by country (Canada, France, UK, USA, other = Australia, Mexico, Spain), sex (female, male), and SSc subtype (diffuse cutaneous, limited cutaneous or sine scleroderma). Only participants with complete data for variables required for classification into strata were included in these analyses.

Healthier individuals in the SPIN Cohort may be less likely to be lost to follow-up.^27^ Thus, we conducted sensitivity analyses that included only participants who completed the itch numerical rating scale for the first time ≤ 10 years after non-RP symptom onset.

### Involvement of People with Lived Experience

Four people with SSc who were members of a SPIN Pain Advisory Team encouraged a study on itch. They formed the SPIN Itch Patient Advisory Team. They advised on a grant proposal, reviewed research questions for this study, and helped interpret results. They are authors. Seven additional people with SSc from SPIN’s Steering Committee who have oversight roles on all SPIN research reviewed the study protocol and the present manuscript and are authors.

## RESULTS

As of October 10, 2020, the last date the itch measure was administered, 2430 participants had enrolled in the SPIN Cohort and completed at least one assessment. Of these, 2365 (97·3%) had completed at least one past-week itch rating, and 2173 of these (91·9%) had time since non-RP symptom onset data and were included. 1896 (87·3%) participants were female, and 1735 (79·8%) were White. Mean age was 54·7 (SD 12·7) years. Participants were from the USA (778 [35·8%]); France (590 [27·2%]); Canada (507 [23·3%]); the UK (191 [8·8%]); and Australia, Mexico, or Spain (107 [4·9%]). Mean time since non-RP symptom onset at enrolment was 11·1 (8·8) years, and 873 (40·2%) participants had diffuse cutaneous SSc. See table 1 for participant characteristics and appendix pp 3-5 for participants by site.

**Table 1.**
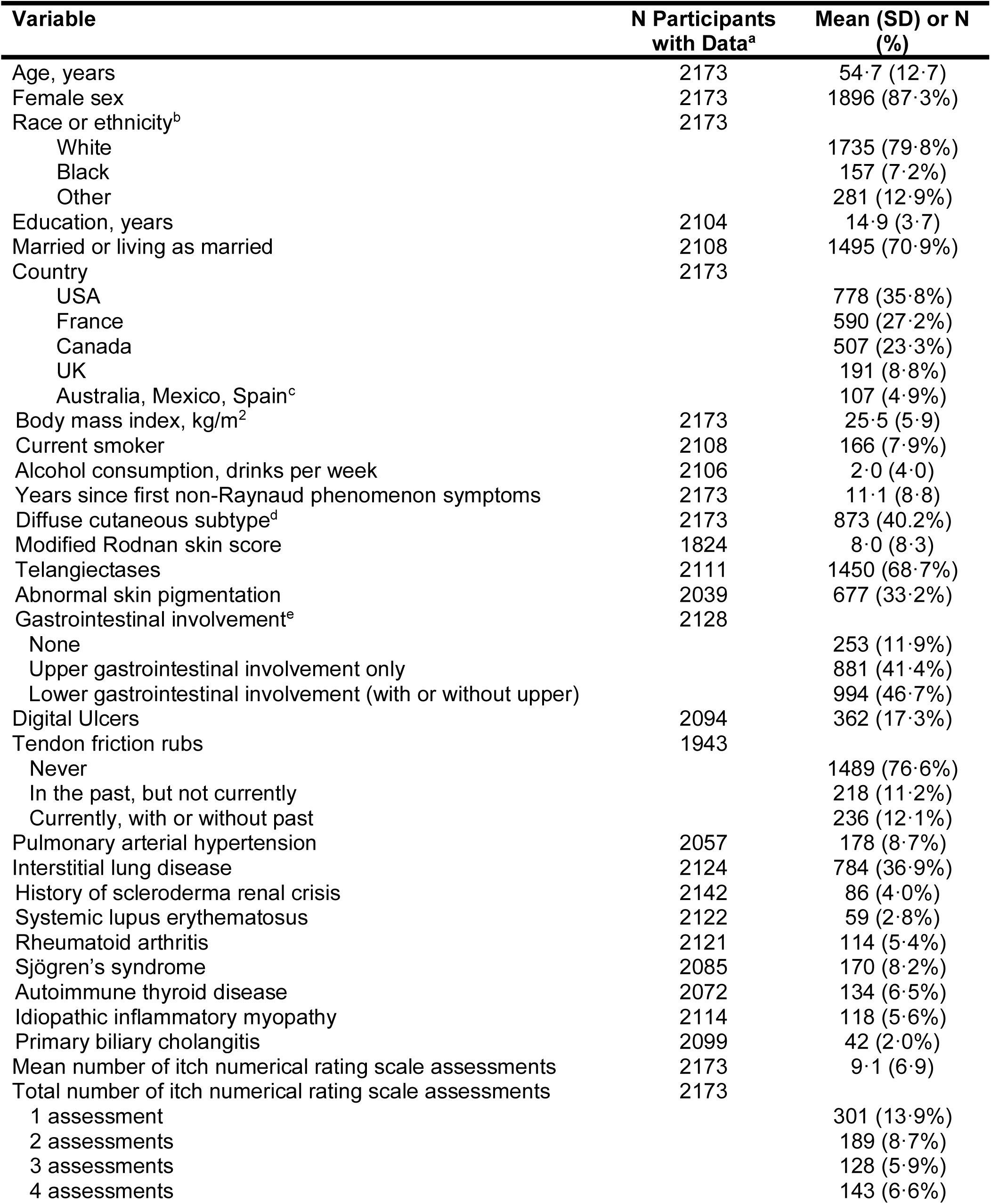
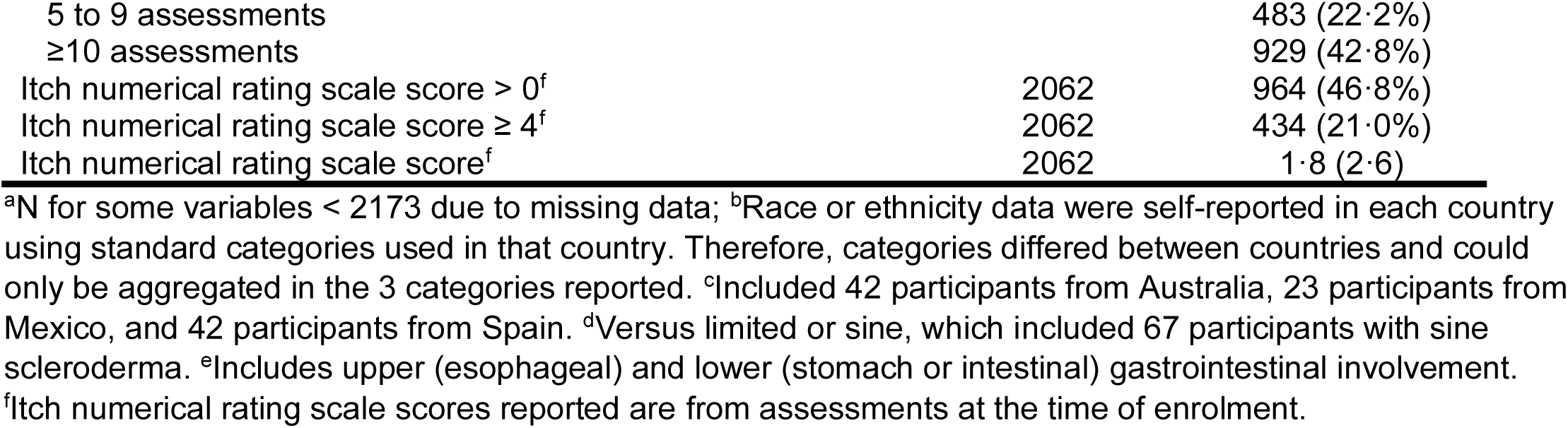
Sociodemographic and disease characteristics of 2173 systemic sclerosis patients at time of SPIN Cohort enrolment.

Participants completed between 1 and 26 past-week itch assessments. 301 (13·9%) participants had one past-week itch severity assessment, 189 (8·7%) had 2, 128 (5·9%) had 3, 143 (6·6%) had 4, 483 (22·2%) had between 5 and 9, and 929 (42·8%) 10 or more assessments. Mean (SD) number of included assessments per participant was 9·1 (6·9). Of 19 733 total assessments included in analyses, 1729 (8·8%) were completed between ages 18 and 39 years, 11 855 (60·1%) between ages 40 and 64 years, 5901 (29·9%) between ages 65 and 79 years, and 248 (1·3%) at ages ≥ 80 years. Figure 1 shows the distribution of assessments by years since non-RP symptom onset. See appendix p 6 for the distribution of assessments by itch severity score.

**Figure 1.**
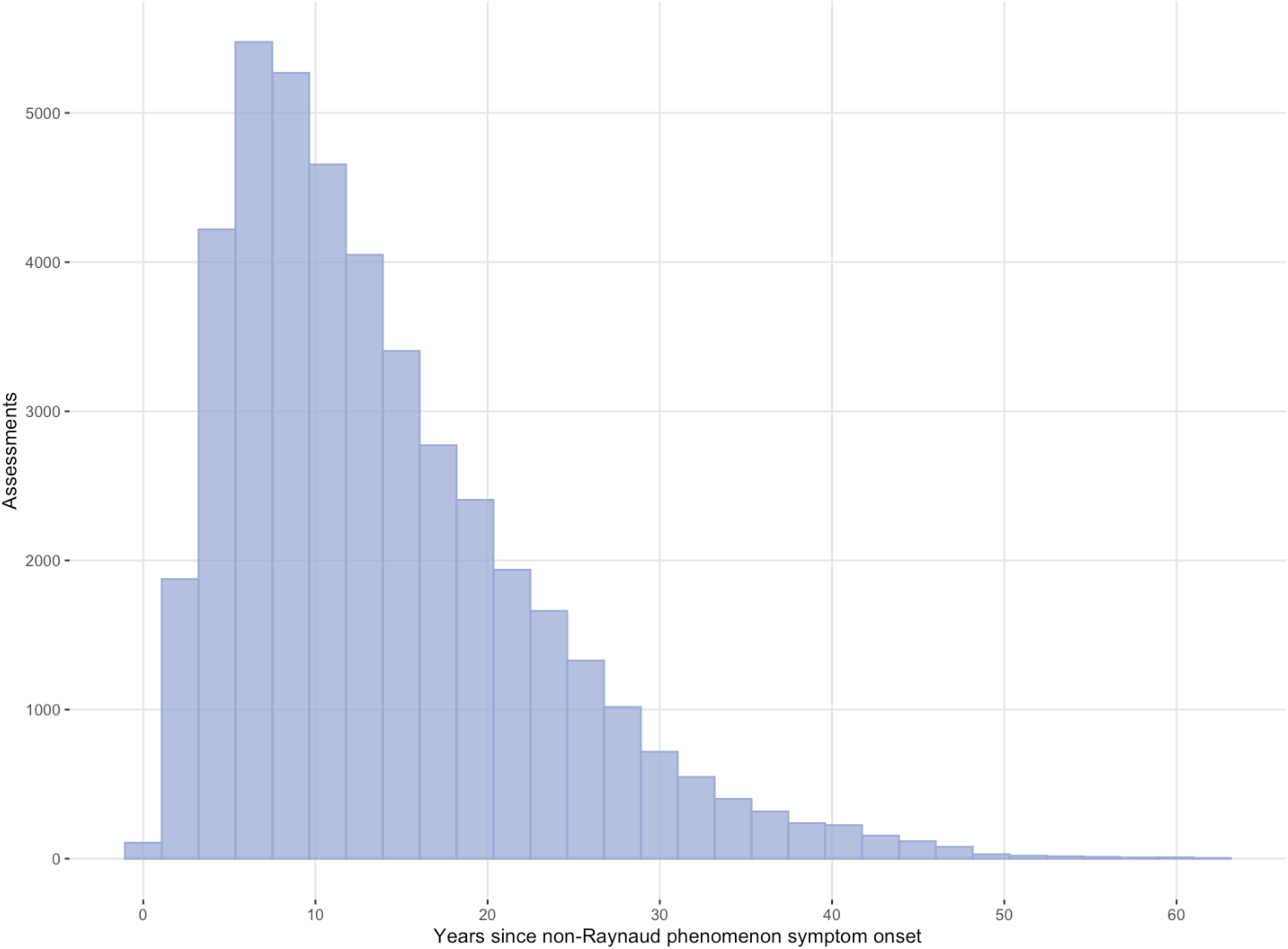
Frequency distribution of the number of itch severity numerical rating scale assessments by time since non-Raynaud phenomenon symptom onset.

Figure 2 shows trajectories of predicted probability of the presence of any itch (numerical rating scale score > 0) and Figure 3 the predicted severity for participants who reported any itch, both across time by age of onset. Separate figures for each age of onset with 95% CIs are shown in the appendix (pp 7-8). Predicted values of probability of the presence of any itch and severity among those who reported any itch are in Table 2. All predicted probabilities of reporting itch were between 35·0% (95% CI 31·8% to 38·5%; age of non-RP symptom onset 20 years, 35 years since onset) and 36·8% (95% CI 33·3% to 40·4%; age of non-RP symptom onset 20 years, 4 years since onset). The lowest predicted severity among participants who reported any itch was 4·1 (95% CI 4·1 to 4·1; age of non-RP symptom onset 50 years, 20 years since onset), and the highest was 4·4 (95% CI 4·3 to 4·4; age of non-RP symptom onset 20 years, 4 years since onset). For all non-RP symptom onset ages, predicted itch severity decreased less than 0·2 points for all 10-year increments (e.g., 5 years to 15 years, 20 years to 30 years) in years since non-RP symptom onset. Predicted values from sensitivity analyses are shown in the appendix (pp 9-11). Results were similar to results from main analyses.

**Figure 2.**
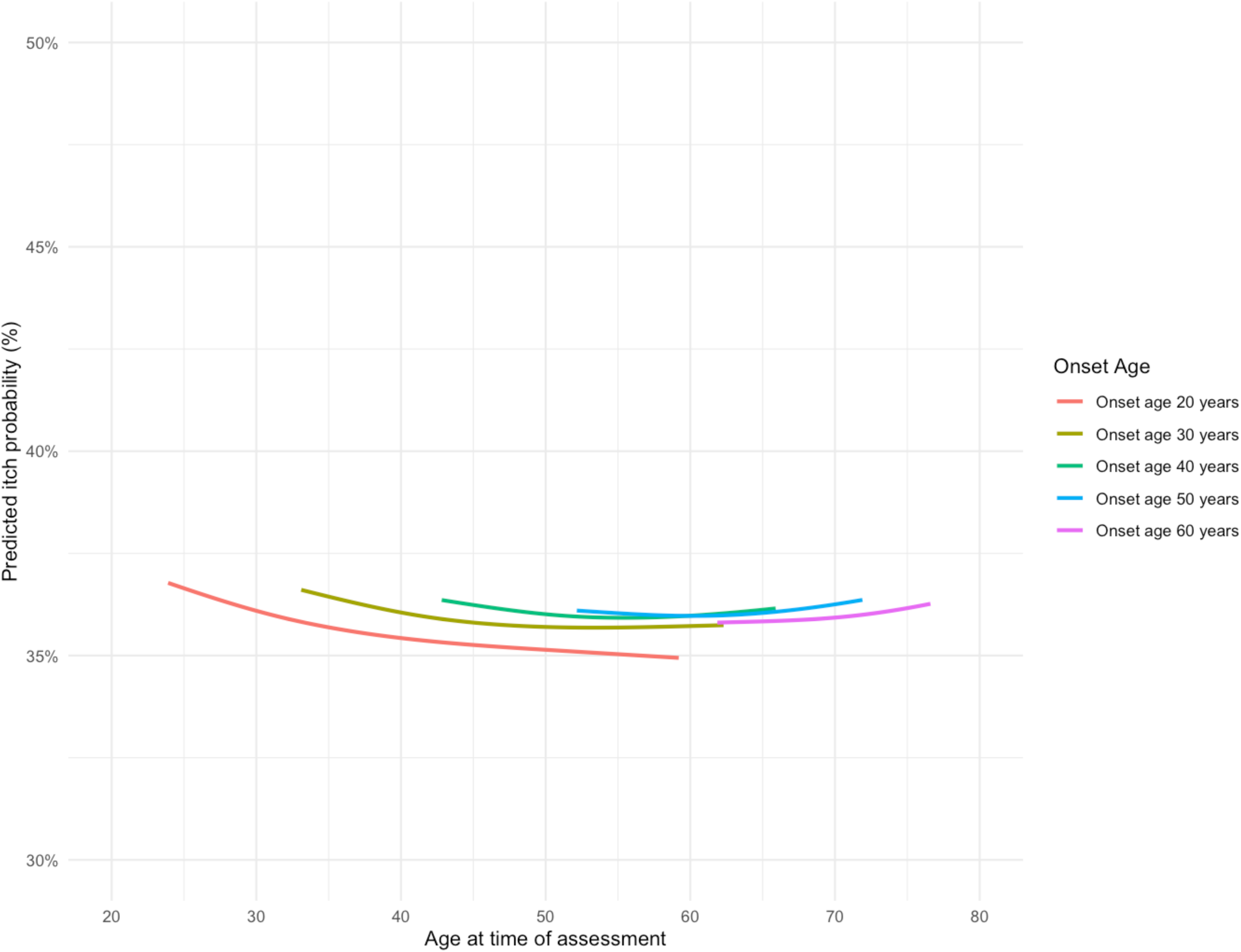
Predicted probability of itch (numerical rating scale score > 0) by age at time of assessment for different ages of non-Raynaud phenomenon symptom onset. For each predicted trajectory (e.g., non-Raynaud phenomenon symptom onset age 30 years), we defined trajectory boundaries using participants with onset ages ± 5 years from the prediction age (e.g., ≥25 to <35 years). We started each trajectory at the 10^th^ percentile of time since non-RP symptom onset and finished at the 90^th^ percentile to ensure adequate data for estimates. Resulting ranges of time since non-Raynaud phenomenon symptom onset in the figure are for onset age 20 years, 3·9 to 39·2 years; onset age 30 years, 3·1 to 32·3 years; onset age 40 years, 2·8 to 25·9 years; onset age 50 years, 2·2 to 21·9 years; and onset age 60 years, 1·9 to 16·6 years.

**Figure 3.**
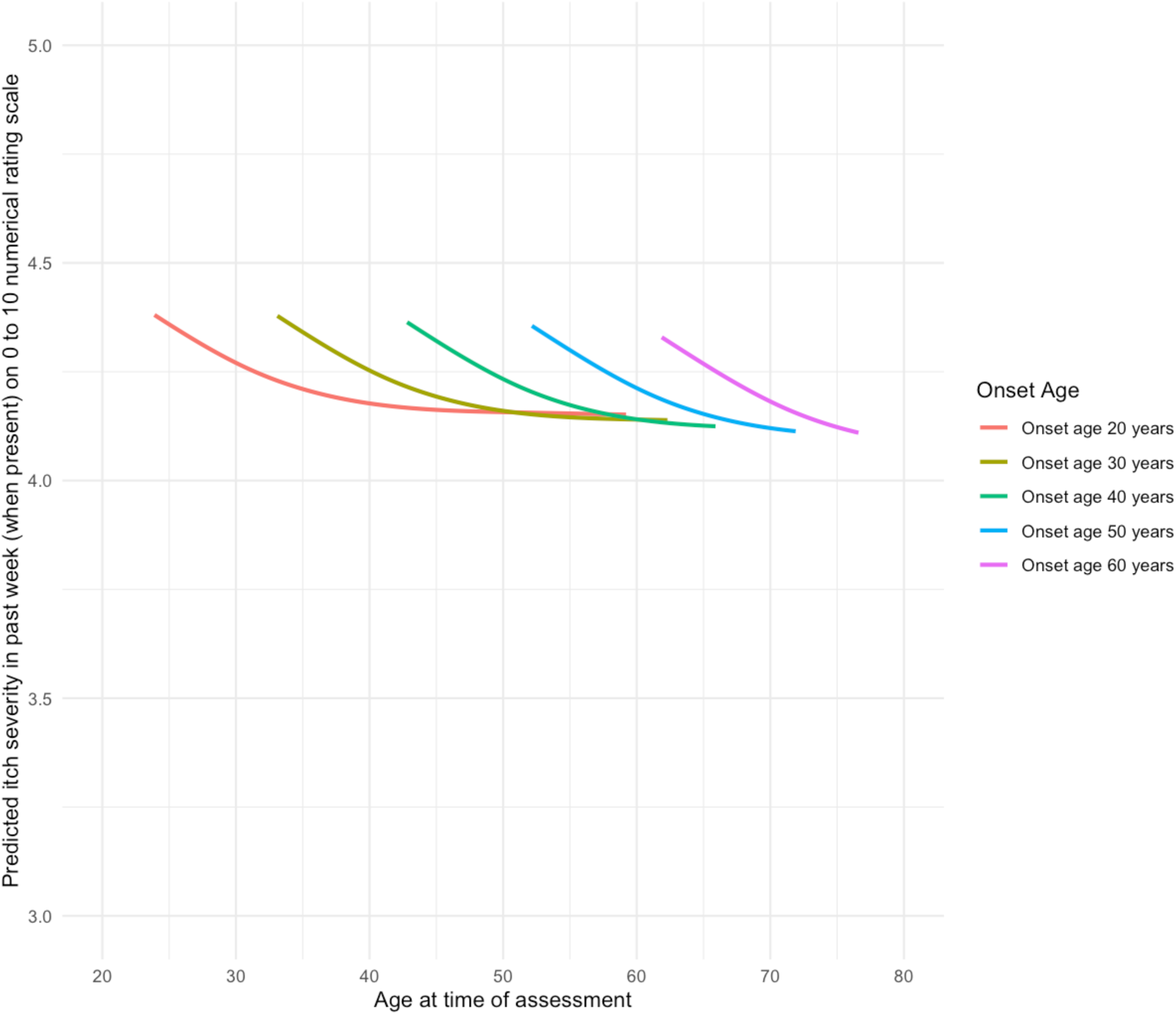
Predicted severity of past-week itch, if present, on 0 to 10 numerical rating scale by age at time of assessment for different ages of non-Raynaud phenomenon symptom onset. For each predicted trajectory (e.g., non-Raynaud phenomenon symptom onset age 30 years), we defined trajectory boundaries using participants with onset ages ± 5 years from the prediction age (e.g., ≥25 to <35 years). We started each trajectory at the 10^th^ percentile of time since non-RP symptom onset and finished at the 90^th^ percentile to ensure adequate data for estimates. Resulting ranges of time since non-Raynaud phenomenon symptom onset in the figure are for onset age 20 years, 3·9 to 39·2 years; onset age 30 years, 3·1 to 32·3 years; onset age 40 years, 2·8 to 25·9 years; onset age 50 years, 2·2 to 21·9 years; and onset age 60 years, 1·9 to 16·6 years.

**Table 2.** Predicted values of probability of itch presence (numerical rating scale score > 0) and itch severity in past week (0 to 10 numerical rating scale), if present, for an average participant at 2 years, 3 years, 4 years, 5 years, 7 years, and at 5-year intervals from 10 years to 35 years post non-Raynaud phenomenon symptom onset with 95% confidence intervals by age at non-Raynaud phenomenon symptom onset.

|  | Age of non-Raynaud phenomenon symptom onset |  |  |  |  |
| --- | --- | --- | --- | --- | --- |
|  | 20 years | 30 years | 40 years | 50 years | 60 years |
| Years since non-Raynaud phenomenon symptom onset<sup>a</sup> | Itch probability (%) |  |  |  |  |
| 2 years | .. | .. | .. | .. | 35.8 (33.5 to 38.2) |
| 3 years | .. | .. | 36.3 (33.9 to 38.9) | 36.1 (34.0 to 38.2) | 35.8 (33.7 to 38.0) |
| 4 years | 36.8 (33.3 to 40.4) | 36.5 (33.7 to 39.4) | 36.3 (34.1 to 38.6) | 36.1 (34.1 to 38.0) | 35.8 (33.8 to 37.9) |
| 5 years | 36.6 (33.4 to 40.0) | 36.4 (33.9 to 39.1) | 36.2 (34.2 to 38.3) | 36.0 (34.2 to 37.9) | 35.8 (33.9 to 37.8) |
| 7 years | 36.4 (33.6 to 39.3) | 36.3 (34.0 to 38.6) | 36.1 (34.3 to 38.0) | 36.0 (34.4 to 37.6) | 35.9 (34.1 to 37.7) |
| 10 years | 36.1 (33.7 to 38.6) | 36.1 (34.1 to 38.1) | 36.0 (34.4 to 37.7) | 36.0 (34.4 to 37.6) | 35.9 (34.2 to 37.7) |
| 15 years | 35.7 (33.3 to 38.1) | 35.8 (33.9 to 37.8) | 35.9 (34.2 to 37.7) | 36.0 (34.3 to 37.8) | 36.2 (34.2 to 38.2) |
| 20 years | 35.4 (33.0 to 37.9) | 35.7 (33.8 to 37.6) | 36.0 (34.3 to 37.7) | 36.3 (34.3 to 38.2) | .. |
| 25 years | 35.3 (32.7 to 37.9) | 35.7 (33.8 to 37.6) | 36.1 (34.2 to 38.1) | .. | .. |
| 30 years | 35.1 (32.4 to 38.0) | 35.7 (33.5 to 38.0) | .. | .. | .. |
| 35 years | 35.0 (31.8 to 38.5) | .. | .. | .. | .. |
| Itch severity in past week (when present) on 0 to 10 numerical rating scale |  |  |  |  |  |
| 2 years | .. | .. | .. | .. | 4.3 (4.3 to 4.5) |
| 3 years | .. | .. | 4.4 (4.3 to 4.4) | 4.3 (4.3 to 4.4) | 4.3 (4.3 to 4.3) |
| 4 years | 4.4 (4.3 to 4.4) | 4.4 (4.3 to 4.4) | 4.3 (4.3 to 4.4) | 4.3 (4.3 to 4.4) | 4.3 (4.3 to 4.3) |
| 5 years | 4.4 (4.3 to 4.4) | 4.3 (4.3 to 4.4) | 4.3 (4.3 to 4.4) | 4.3 (4.3 to 4.3) | 4.3 (4.2 to 4.3) |
| 7 years | 4.3 (4.3 to 4.4) | 4.3 (4.3 to 4.3) | 4.3 (4.3 to 4.3) | 4.3 (4.2 to 4.3) | 4.2 (4.2 to 4.2) |
| 10 years | 4.3 (4.2 to 4.3) | 4.3 (4.2 to 4.3) | 4.2 (4.2 to 4.3) | 4.2 (4.2 to 4.2) | 4.2 (4.1 to 4.2) |
| 15 years | 4.2 (4.2 to 4.2) | 4.2 (4.2 to 4.2) | 4.2 (4.2 to 4.2) | 4.2 (4.1 to 4.2) | 4.1 (4.1 to 4.1) |
| 20 years | 4·2 (4·2 to 4·2) | 4·2 (4·1 to 4·2) | 4·1 (4·1 to 4·2) | 4·1 (4·1 to 4·1) | ·· |
| 25 years | 4·2 (4·1 to 4·2) | 4·1 (4·1 to 4·2) | 4·1 (4·1 to 4·2) | ·· | ·· |
| 30 years | 4·2 (4·1 to 4·2) | 4·1 (4·1 to 4·2) | ·· | ·· | ·· |
| 35 years | 4·2 (4·1 to 4·2) | ·· | ·· | ·· | ·· |

<sup>a</sup>For each non-Raynaud phenomenon symptom onset age (e.g., 30 years), we defined boundaries using participants with onset ages $\pm 5$ years from the prediction age (e.g., $\geq 25$ to $< 35$ years). We reported predicted values from the 10<sup>th</sup> percentile of years since non-Raynaud phenomenon symptom onset at the first assessment to the 90<sup>th</sup> percentile at the last assessment to ensure adequate data for estimates.

Regression parameters for models in main and sensitivity analyses are shown in appendices (pp 12-13). There was a statistically significant interaction between age of onset and time since onset for itch presence (appendix p 12) but not itch severity (appendix p 13). However, as can be seen in Figure 3, the magnitude of the interaction for itch presence was negligible.

There was no significant association between sex and itch presence or severity (appendix pp 12-13). Compared to participants from the USA, participants from all other countries had higher odds of itch presence (odds ratios 1·34 to·1.78), and participants from the UK had statistically significantly, though minimally (about 3% difference or 0·1 points), higher itch severity when present (appendix pp 12-13). There was a significant association between diffuse cutaneous SSc and higher odds of itch presence (odds ratio 1·25 [95% CI 1·16 to 1·34]; appendix p 12). Figures by sex, country, and disease subtype are shown in appendices pp 14-19.

## DISCUSSION

We analysed 19 733 itch assessments from 2173 participants from 7 countries and simultaneously modelled probability of having any itch and, if present, itch severity. We accounted for both normal aging and SSc disease duration by including age of onset of non-RP symptoms and time since onset in our models. Contrary to common assumptions,^5,10^ itch did not diminish following early disease or worsen over time. Prevalence and mean severity appear to be stable.

Predicted past-week itch presence probabilities for all onset ages and years since onset were between 35% and 37%. When itch was present, predicted severity on a 0 to 10 scale was between 4·1 and 4·4 points, a moderate level,^18^ for all onset ages and times since onset. For all onset ages, predicted itch severity decreased by < 0·2 points for all 10-year differences (e.g., 5 years to 15 years) in time since non-RP symptom onset. Due to the large number of assessments evaluated, we detected a statistically significant interaction between age of onset and time since onset for itch presence and statistically significant differences between countries. All magnitudes, however, were negligible (e.g., about 0·1 to 0·4 points) compared to the itch numerical rating scale minimal important difference (3 to 4 points).^20^

No previous longitudinal studies have evaluated itch presence or severity in SSc. Our findings are generally consistent with several cross-sectional studies that did not find a statistically significant association between disease duration and itch in SSc.^3,7,11,12^ Cross-sectional studies have consistently found the prevalence of itch to be higher among older adults in the general population.^8,9^ The largest study, a 2024 study of 50 552 participants from 20 countries found that past-week itch was present among 23% of people without skin disease aged 65 years or older compared to 20% among people aged 40 to 64 years and 16% among people aged 16 to 39 years; among people with skin disease, 66% of those with a skin condition aged ≥ 65 reported the presence of itch compared to 61% of those aged 40 to 64 years and 50% among those aged 16 to 39 years.^8^ A 2022 systematic review (17 studies, 28 666 participants) found that, among older adults, prevalence may increase from about 12% among those aged 60 to 69 years to > 50% among those aged 80 years or older.^9^

Findings from general population studies raise the question of whether increased itch related to normal aging may be masking decreases in itch in later stage disease in SSc. Our results are not consistent with that idea. We found that change in itch prevalence and severity was minimal regardless of age of disease onset. Furthermore, the estimated prevalence of itch of 35% to 37% across ages of onset and time since onset in our study is substantially higher than all age-group estimates, except for people aged 80 and older and people with diagnosed skin conditions.

Burden from itch in SSc appears to be similar to other systemic diseases where itch is understood to be a common disease feature that is researched and managed as part of routine clinical care. Moderate-to-severe itch, usually defined as a score of 4 or greater on a 0-10 scale, is reported by approximately 20% to 50% of patients with chronic kidney disease^28^ or chronic liver disease.^29^ In our study, 21% of participants scored 4 or greater at the time of enrolment in the SPIN Cohort. Itch, however, is rarely studied and relatively little is known about itch in SSc compared with chronic kidney disease and chronic liver disease. Research on itch in SSc is needed to better understand its pathogenesis and to develop and test management strategies.^5^ The pathophysiology of itch in SSc remains poorly defined, but accumulating evidence suggests that cutaneous fibrosis, microvascular injury, and neuro-immune dysregulation—potentially involving altered small-fiber nerve architecture and pruritogenic cytokines—may promote peripheral sensory sensitisation and chronic pruritus.^30^

Strengths of our study include the large multinational cohort with more than 19 000 itch severity assessments from >2100 included participants from 49 sites in 7 countries. The number of assessments was over 20 times larger than the number of assessments in the next largest study that evaluated itch in SSc.^6^ Our use of zero-inflated modelling allowed us to simultaneously evaluate itch presence and itch severity when present, compared to previous studies which only considered whether itch was present.^3,6,11,12^ The modelling strategy that we used also allowed us to take into consideration components of chronological age and time since SSc onset and to consider non-linear associations between these variables and itch.

There are also limitations to consider. First, SPIN Cohort participants are recruited from specialty SSc centres in highly developed countries and participation requires answering questions via online forms, both of which might reduce generalizability of results. Second, we did not have time-varying data on factors that may contribute to itch trajectories, although the purpose of this study was to map trajectories over time and not to model putative causal factors. Third, we studied aggregate probability of experiencing itch and itch severity, which does not elucidate individual trajectory patterns.

We believe that our study is the largest in any population to evaluate itch presence and severity longitudinally over multiple years. It is the first longitudinal study of itch in SSc. We found that itch was present in 35% to 37% of people with SSc across all combinations of ages of onset and disease duration. Among people with itch, mean severity level was consistently between 4·1 and 4·4 points on a 0-10 scale, a moderate level,^18^ for all age and duration combinations. Our findings highlight that, contrary to assumptions, itch is a common and salient feature of SSc across the course of the disease. Research is needed to examine patterns of itch to determine how they may vary between individuals, advance understandings of itch pathology, and test management approaches. Itch among people with SSc should be evaluated and treated as part of routine care.

## Author Contributions

MG, CDS, EN, LMisery, JLM, TM, SR, MS, AP, JP, GY, AB, and BDT contributed to the conception and design of the study. TF, SH, JP, BC, LKH, MDM, and LMisery contributed to participant enrolment and collection of medical data. MEC, LK, and BDT contributed to data collection via SPIN Cohort management. MEC, LK, SH, MS, SJB, BC, CF, AG, KG, GG, LKH, ALJ, VLM, MDM, YP, MR, JS, RKW, LMouthon, and BDT contributed to oversight and management of the SPIN Cohort via roles on the SPIN Steering Committee. MG, AB, and BDT contributed to development of the statistical analysis plan or data analysis; MG and BDT accessed and verified the underlying data. All authors contributed to interpretation of results. MG and BDT drafted the manuscript. All authors provided a critical review and approved the final manuscript. BDT is the guarantor.

## Data Sharing

De-identified participant data with a data dictionary and analysis codes that were used to generate the results reported in this article will be made available upon request to the corresponding author and presentation of a methodologically sound proposal that is approved by the Scleroderma Patient-centred Intervention Network Data Access and Publications Committee. Data will be available beginning 12 months after publication. Data requesters will need to sign a data transfer agreement.

## Acknowledgements

Funding for the study was provided by a Skin Investigation Network of Canada Team Development Award. The Scleroderma Patient-centered Intervention Network (SPIN) Cohort has received funding from: the Canadian Institutes of Health Research (TR3-119192; PJT-149073; PJT-148504; PJT-195879; PJT-203755); the Arthritis Society; the Lady Davis Institute for Medical Research of the Jewish General Hospital, Montréal, Québec, Canada; the Jewish General Hospital Foundation, Montréal, Québec, Canada; McGill University, Montréal, Québec, Canada; the Scleroderma Society of Ontario; Scleroderma Canada; Sclérodermie Québec; Scleroderma Manitoba; Scleroderma Atlantic; the Scleroderma Association of BC; Scleroderma SASK; Scleroderma Australia; Scleroderma New South Wales; Scleroderma Victoria, and the Scleroderma Foundation of California. Ms. Dal Santo was supported by a Fonds de Recherche du Québec—Société et Culture Doctoral Research Award and Dr. Thombs by a Tier 1 Canada Research Chair, both outside of the present work. No sponsor or funder was involved in the study design; in the collection, analysis, and interpretation of the data; in the writing of the report; or in the decision to submit the paper for publication.

SPIN Investigators include: Claire Adams, Lady Davis Institute of the Jewish General Hospital, Montréal, Québec, Canada; Vanessa Cook, Scleroderma Society of Ontario and Scleroderma Canada, Hamilton, Ontario, Canada; Marie Hudson, Department of Medicine, McGill University, Montréal, Québec, Canada; Yanne Perriault, Association des Sclérodermiques de France, Baccon, France; Christian Agard, Centre Hospitalier Universitaire - Hôtel-Dieu de Nantes, Nantes, France; Laurent Alric, CHU Rangueil, Toulouse, France; Marc André, Centre Hospitalier Universitaire Gabriel-Montpied, Clermont-Ferrand, France; Floryan Beaslay, CHU La Réunion, Saint-Denis, La Réunion, France; Elana J. Bernstein, Columbia University, New York, New York, United States; Sabine Berthier, Centre Hospitalier Universitaire Dijon Bourgogne, Dijon, France; Lyne Bissonnette, Université de Sherbrooke, Sherbrooke, Québec, Canada; Sophie Blaise, CHU Grenoble Alpes, Grenoble, France; Alessandra Bruns, Université de Sherbrooke, Sherbrooke, Québec, Canada; Carlotta Cacciatore, Assistance Publique - Hôpitaux de Paris, Hôpital St-Louis, Paris, France; Patricia Carreira, Servicio de Reumatologia del Hospital 12 de Octubre, Madrid, Spain; Marion Casadevall, Assistance Publique - Hôpitaux de Paris, Hôpital Cochin, Paris, France; Lorinda Chung, Stanford University, Stanford, California, United States; Benjamin Crichi, Assistance Publique - Hôpitaux de Paris, Hôpital St-Louis, Paris, France; Thylbert Deltombe, CHU La Réunion, Saint-Denis, La Réunion, France; Christopher P. Denton, Royal Free London Hospital, London, United Kingdom; Tannvir Desroche, CHU La Réunion, Saint-Denis, La Réunion, France; Robyn Domsic, University of Pittsburgh, Pittsburgh, Pennsylvania, United States; James V. Dunne, St. Paul’s Hospital and University of British Columbia, Vancouver, British Columbia, Canada; Bertrand Dunogue, Assistance Publique - Hôpitaux de Paris, Hôpital Cochin, Paris, France; Regina Fare, Servicio de Reumatologia del Hospital 12 de Octubre, Madrid, Spain; Dominique Farge-Bancel, Assistance Publique - Hôpitaux de Paris, Hôpital St-Louis, Paris, France; Paul R. Fortin, CHU de Québec - Université Laval, Québec, Québec, Canada; Loraine Gauzère, CHU La Réunion, Saint-Denis, La Réunion, France; Anne Gerber, CHU La Réunion, Saint-Denis, La Réunion, France; Jessica K. Gordon, Hospital for Special Surgery, New York City, New York, United States; Brigitte Granel-Rey, Université, and Assistance Publique - Hôpitaux de Marseille, Hôpital Nord, Marseille, France; Aurélien Guffroy, Les Hôpitaux Universitaires de Strasbourg, Nouvel Hôpital Civil, Strasbourg, France; Geneviève Gyger, Jewish General Hospital and McGill University, Montréal, Québec, Canada; Eric Hachulla, Centre Hospitalier Régional Universitaire de Lille, Hôpital Claude Huriez, Lille, France; Daphna Harel, New York University, New York, New York, United States; Monique Hinchcliff, Yale School of Medicine, New Haven, Connecticut, United States; Michael Hugues, Salford Royal NHS Foundation Trust, Salford, United Kingdom; Alena Ikic, CHU de Québec - Université Laval, Québec, Québec; Sindhu R. Johnson, Toronto Scleroderma Program, Mount Sinai Hospital, Toronto Western Hospital, and University of Toronto, Toronto, Ontario, Canada; Nader Khalidi, McMaster University, Hamilton, Ontario, Canada; Kimberly S. Lakin, Hospital for Special Surgery, New York City, New York, United States; Marc Lambert, Centre Hospitalier Régional Universitaire de Lille, Hôpital Claude Huriez, Lille, France; Maggie Larche, University of Calgary, Calgary, Alberta, Canada; David Launay, Centre Hospitalier Régional Universitaire de Lille, Hôpital Claude Huriez, Lille, France; Yvonne C. Lee, Northwestern University, Chicago, Illinois, United States; Paul Legendre, Centre Hospitalier du Mans, Le Mans, France; Catarina Leite, University of Minho, Braga, Portugal; Hélène Maillard, Centre Hospitalier Régional Universitaire de Lille, Hôpital Claude Huriez, Lille, France; Nancy Maltez, University of Ottawa, Ottawa, Ontario, Canada; Joanne Manning, Salford Royal NHS Foundation Trust, Salford, United Kingdom; Isabelle Marie, CHU Rouen, Hôpital de Bois-Guillaume, Rouen, France; Maria Martin Lopez, Servicio de Reumatologia del Hospital 12 de Octubre, Madrid, Spain; Thierry Martin, Les Hôpitaux Universitaires de Strasbourg, Nouvel Hôpital Civil, Strasbourg, France; Ariel Masetto, Université de Sherbrooke, Sherbrooke, Québec, Canada; Arsène Mekinian, Assistance Publique - Hôpitaux de Paris, Hôpital St-Antoine, Paris, France; Sheila Melchor Díaz, Servicio de Reumatologia del Hospital 12 de Octubre, Madrid, Spain; Morgane Mourguet, CHU Rangueil, Toulouse, France; Christelle Nguyen, Université Paris Descartes, Université de Paris, Paris, France, and Assistance Publique - Hôpitaux de Paris, Paris, France; Karen Nielsen, Scleroderma Society of Ontario, Hamilton, Ontario, Canada; Mandana Nikpour, St Vincent’s Hospital and University of Melbourne, Melbourne, Victoria, Australia; Louis Olagne, Centre Hospitalier Universitaire Gabriel-Montpied, Clermont-Ferrand, France; Vincent Poindron, Les Hôpitaux Universitaires de Strasbourg, Nouvel Hôpital Civil, Strasbourg, France; Susanna Proudman, Royal Adelaide Hospital and University of Adelaide, Adelaide, South Australia, Australia; Grégory Pugnet, CHU Rangueil, Toulouse, France; Loïc Raffray, CHU La Réunion, Saint-Denis, La Réunion, France; François Rannou, Université Paris Descartes, Université de Paris, Paris, France, and Assistance Publique - Hôpitaux de Paris, Paris, France; Alexis Régent, Assistance Publique - Hôpitaux de Paris, Hôpital Cochin, Paris, France; Frederic Renou, CHU La Réunion, Saint-Denis, La Réunion, France; Sébastien Rivière, Assistance Publique - Hôpitaux de Paris, Hôpital St-Antoine, Paris, France; David Robinson, University of Manitoba, Winnipeg, Manitoba, Canada; Esther Rodríguez Almazar, Servicio de Reumatologia del Hospital 12 de Octubre, Madrid, Spain; Tatiana Sofia Rodríguez-Reyna, Instituto Nacional de Ciencias Médicas y Nutrición Salvador Zubirán, Mexico City, Mexico; Sophie Roux, Université de Sherbrooke, Sherbrooke, Québec, Canada; Perrine Smets, Centre Hospitalier Universitaire Gabriel-Montpied, Clermont-Ferrand, France; Vincent Sobanski, Centre Hospitalier Régional Universitaire de Lille, Hôpital Claude Huriez, Lille, France; Robert F. Spiera, Hospital for Special Surgery, New York City, New York, United States; Virginia Steen, Georgetown University, Washington, DC, United States; Evelyn Sutton, Dalhousie University, Halifax, Nova Scotia, Canada; Carter Thorne, Southlake Regional Health Centre, Newmarket, Ontario, Canada; Damien Vagner, CHU La Réunion, Saint-Denis, La Réunion, France; John Varga, Division of Rheumatology, Department of Internal Medicine, University of Michigan, Ann Arbor, Michigan, United States; Pearce Wilcox, St. Paul’s Hospital and University of British Columbia, Vancouver, British Columbia, Canada; Djellza Dani, Jewish General Hospital, Montréal, Québec, Canada; Monica D’Onofrio, Jewish General Hospital, Montréal, Québec, Canada; Sophie Hu, Jewish General Hospital, Montréal, Québec; Nicole Iordache, Jewish General Hospital, Montréal, Québec, Canada; Elsa-Lynn Nassar, Jewish General Hospital, Montréal, Québec, Canada; and Maria Nassar, Jewish General Hospital, Montréal, Québec, Canada.

## REFERENCES

1 Denton CP, Khanna D. Systemic sclerosis. Lancet 2017;390(10103):1685–99. doi:10.1016/S0140-6736(17)30933-9

2 Bassel M, Hudson M, Taillefer SS, Schieir O, Baron M, Thombs BD. Frequency and impact of symptoms experienced by patients with systemic sclerosis: results from a Canadian National Survey. Rheumatology 2011;50(4):762–7. doi:10.1093/rheumatology/keq310

3 Razykov I, Levis B, Hudson M, Baron M, Thombs BD; Canadian Scleroderma Research Group. Prevalence and clinical correlates of pruritus in patients with systemic sclerosis: an updated analysis of 959 patients. Rheumatology 2013;52(11):2056–61. doi:10.1093/rheumatology/ket275

4 El-Baalbaki G, Razykov I, Hudson M, et al. Association of pruritus with quality of life and disability in systemic sclerosis. Arthritis Care Res 2010;62(10):1489–95. doi:10.1002/acr.20257

5 Frech TM, Baron M. Understanding itch in systemic sclerosis in order to improve patient quality of life. Clin Exp Rheumatol 2013;31(2 Suppl 76):81–8.

6 Razykov I, Thombs BD, Hudson M, Bassel M, Baron M; Canadian Scleroderma Research Group. Prevalence and clinical correlates of pruritus in patients with systemic sclerosis. Arthritis Rheum 2009;61(12):1765–70. doi:10.1002/art.25010

7 Samela T, Raimondi G, Cordella G, et al. Living with systemic sclerosis: prevalence of itch and other skin symptoms and their association with psychosocial issues. J Health Psychol 2026;31(2):867–81. doi:10.1177/13591053251342549

8 Yosipovitch G, Skayem C, Saint Aroman M, et al. International study on prevalence of itch: examining the role of itch as a major global public health problem. Br J Dermatol 2024;191(5):713–8. doi:10.1093/bjd/ljae260

9 Chen S, Zhou F, Xiong Y. Prevalence and risk factors of senile pruritus: a systematic review and meta-analysis. BMJ Open 2022;12(2):e051694. doi:10.1136/bmjopen-2021-051694

10 Herrick AL, Assassi S, Denton CP. Skin involvement in early diffuse cutaneous systemic sclerosis: an unmet clinical need. Nat Rev Rheumatol 2022;18(5):276–85. doi:10.1038/s41584-022-00765-9

11 Gourier G, Théréné C, Mazeas M, et al. Clinical characteristics of pruritus in systemic sclerosis vary according to the autoimmune subtype. Acta Derm Venereol 2018;98(8):735–41. doi:10.2340/00015555-2980

12 Stull CM, Weaver LA, Valdes-Rodriguez R, et al. Characteristics of chronic itch in systemic sclerosis: a cross-sectional survey. Acta Derm Venereol 2018;98(8):793–4. doi:10.2340/00015555-2966

13 Dougherty DH, Kwakkenbos L, Carrier ME, et al. The Scleroderma Patient-Centered Intervention Network Cohort: baseline clinical features and comparison with other large scleroderma cohorts. Rheumatology 2018;57(9):1623–31. doi:10.1093/rheumatology/key139

14 Vandenbroucke JP, von Elm E, Altman DG, et al. Strengthening the Reporting of Observational Studies in Epidemiology (STROBE): explanation and elaboration. Int J Surg 2014;12(12):1500–24. doi:10.1016/j.ijsu.2014.07.014

15 Isaacson RA. Text recycling in scientific writing: what editors need to know. Sci Ed 2023;46(3):107–9. doi:10.36591/SE-D-4603-10

16 van den Hoogen F, Khanna D, Fransen J, et al. 2013 classification criteria for systemic sclerosis: an American college of rheumatology/European league against rheumatism collaborative initiative. Ann Rheum Dis 2013;72(11):1747–55. doi:10.1136/annrheumdis-2013-204424

17 Clements P, Lachenbruch P, Siebold J, et al. Inter and intraobserver variability of total skin thickness score (modified Rodnan TSS) in systemic sclerosis. J Rheumatol 1995;22(7):1281–5.

18 Yosipovitch G, Reaney M, Mastey V, et al. Peak Pruritus Numerical Rating Scale: psychometric validation and responder definition for assessing itch in moderate-to-severe atopic dermatitis. Br J Dermatol 2019;181(4):761–9. doi:10.1111/bjd.17744

19 Phan NQ, Blome C, Fritz F, et al. Assessment of pruritus intensity: prospective study on validity and reliability of the visual analogue scale, numerical rating scale and verbal rating scale in 471 patients with chronic pruritus. Acta Derm Venereol 2012;92(5):502–7. doi:10.2340/00015555-1246

20 Erickson S, Kim BS. Research techniques made simple: itch measurement in clinical trials. J Invest Dermatol 2019;139(2):264–9.e1. doi:10.1016/j.jid.2018.12.004

21 Rizopoulos D. GLMMadaptive: generalized linear mixed models using adaptive Gaussian quadrature [Internet]. GLMMadaptive; 2023 Oct 17 [cited 2024 Dec 24]. Available from: https://drizopoulos.github.io/GLMMadaptive/

22 Rizopoulos D. Zero-inflated and two-part mixed effects models [Internet]. GLMMadaptive; 2023 Oct 17 [cited 2024 Dec 24]. Available from : https://drizopoulos.github.io/GLMMadaptive/articles/ZeroInflated_and_TwoPart_Models.html#two-part-mixed-effects-model-for-semi-continuous-data

23 Neelon B, O’Malley AJ. Two-part models for zero-modified count and semicontinuous data. In: Sobolev B, Gatsonis C, editors. Methods in health services research. New York: Springer; 2017. p. 1–23. doi:10.1007/978-1-4939-6704-9_17-1

24 Griswold ME, Glymour MM. Time and age as longitudinal timescales: multiple useful models are illuminating. Epidemiology 2025;36(4):572–9. doi:10.1097/EDE.0000000000001869

25 Perperoglou A, Sauerbrei W, Abrahamowicz M, Schmid M. A review of spline function procedures in R. BMC Med Res Methodol 2019;19(1):46. doi:10.1186/s12874-019-0666-3

26 Desquilbet L, Mariotti F. Dose-response analyses using restricted cubic spline functions in public health research. Stat Med 2010;29(9):1037–57. doi:10.1002/sim.3841

27 Tong CYM, Koh RYV, Lee ES. A scoping review on the factors associated with the lost to follow-up (LTFU) amongst patients with chronic disease in ambulatory care of high-income countries (HIC). BMC Health Serv Res 2023;23(1):883. doi:10.1186/s12913-023-09863-0

28 Lanot A, Bataille S, Rostoker G, et al. Moderate-to-severe pruritus in untreated or non-responsive hemodialysis patients: results of the French prospective multicenter observational study Pruripreva. Clin Kidney J 2023;16(7):1102–12. doi:10.1093/ckj/sfad032

29 Gungabissoon U, Hunnicutt J, McDermott EJ, et al. Pruritus and health-related quality of life in chronic liver disease: a longitudinal, survey-based cohort study. BMJ Open Gastroenterol 2025;12(1):e001809. doi:10.1136/bmjgast-2025-001809

30 Haber JS, Valdes-Rodriguez R, Yosipovitch G. Chronic pruritus and connective tissue disorders: review, gaps, and future directions. Am J Clin Dermatol 2016;17(5):445– 9. doi:10.1007/s40257-016-0201-9

