## Supplemental Files for "Course of Itch from Systemic Sclerosis Onset: a Scleroderma Patient-Centred Intervention Network Cohort Longitudinal Study"

**APPENDIX**

**Appendix Table 1.** Participants by country and recruitment site (pp 3-5).

**Appendix Figure 1.** Frequency distribution of the number of itch severity numerical rating scale assessments by past-week itch severity score (p 6).

**Appendix Figure 2.** Predicted probability of itch (numerical rating scale score > 0) by age at time of assessment for different ages of non-Raynaud phenomenon symptom onset, with 95% confidence intervals (p 7).

**Appendix Figure 3.** Predicted severity of past-week itch, if present, on 0 to 10 numerical rating scale by age at time of assessment for different ages of non-Raynaud phenomenon symptom onset, with 95% confidence intervals (p 8).

**Appendix Figure 4.** Predicted probability of itch (numerical rating scale score > 0) by age at time of assessment for different ages of non-Raynaud phenomenon symptom onset, including only participants who completed the itch numerical rating scale for the first time ≤ 10 years after non-Raynaud phenomenon symptom onset (p 9).

**Appendix Figure 5.** Predicted severity of past-week itch, if present, on 0 to 10 numerical rating scale by age at time of assessment for different ages of non-Raynaud phenomenon symptom onset, including only participants who completed the itch numerical rating scale for the first time ≤ 10 years after non-Raynaud phenomenon symptom onset (p 10).

**Appendix Table 2.** Predicted values of itch probability (numerical rating scale score > 0) and itch severity in past week (0 to 10 numerical rating scale), if present, for an average participant at 2 years, 3 years, 4 years, 5 years, 7 years and 10 years post non-Raynaud phenomenon symptom onset with 95% confidence intervals by age at non-Raynaud phenomenon symptom onset, including only participants who completed the itch numerical rating scale for the first time ≤ 10 years after non-Raynaud phenomenon symptom onset (p 11).

**Appendix Table 3.** Predictors of itch presence (numerical rating scale score > 0) with 95% confidence intervals from main analysis (N = 2173 participants, 19733 assessments) and sensitivity analysis including only participants who completed the itch numerical rating scale for the first time ≤ 10 years after non-Raynaud phenomenon symptom onset (N = 1192 participants, 10549 assessments) (p 12).

**Appendix Table 4.** Predictors of past-week itch severity, if present, on 0 to 10 numerical rating scale with 95% confidence intervals from main analysis (N = 2173 participants, 19733 assessments) and sensitivity analysis including only participants who completed the itch numerical rating scale for the first time ≤ 10 years after non-Raynaud phenomenon symptom onset (N = 1192 participants, 10549 assessments) (p 13).

**Appendix Figure 6.** Predicted probability of itch (numerical rating scale score > 0) by age at time of assessment for different ages of non-Raynaud phenomenon symptom onset, by sex (p 14).

**Appendix Figure 7.** Predicted severity of past-week itch, if present, on 0 to 10 numerical rating scale by age at time of assessment for different ages of non-Raynaud phenomenon symptom onset, by sex (p 15).

**Appendix Figure 8.** Predicted probability of itch (numerical rating scale score > 0) by age at time of assessment for different ages of non-Raynaud phenomenon symptom onset, by country (p 16).

**Appendix Figure 9.** Predicted severity of past-week itch, if present, on 0 to 10 numerical rating scale by age at time of assessment for different ages of non-Raynaud phenomenon symptom onset, by country (p 17).

**Appendix Figure 10.** Predicted probability of itch (numerical rating scale score > 0) by age at time of assessment for different ages of non-Raynaud phenomenon symptom onset, by disease subtype (p 18).

**Appendix Figure 11.** Predicted severity of past-week itch, if present, on 0 to 10 numerical rating scale by age at time of assessment for different ages of non-Raynaud phenomenon symptom onset, by disease subtype (p 19).

**Appendix Table 1.** Participants by country and recruitment site.

| Site | City | Province, Region, State, or Country | N Included |
|---|---|---|---|
| **Australia** | | | **42** |
| Royal Adelaide Hospital | Adelaide | South Australia | 41 |
| St Vincent's Hospital | Melbourne | Victoria | 1 |
| **Canada** | | | **507** |
| Arthritis Centre, University of Manitoba | Winnipeg | Manitoba | 19 |
| Arthritis Program Research Group (Southlake Regional Health Centre) | Newmarket | Ontario | 16 |
| Capital District Health Authority | Halifax | Nova Scotia | 3 |
| Centre de recherche du CHU de Québec | Québec | Québec | 13 |
| Centre Hospitalier de l'université de Montréal | Montréal | Québec | 1 |
| Centre Hospitalier Universitaire de Sherbrooke | Sherbrooke | Québec | 52 |
| Horizon Health Network | Moncton | New Brunswick | 3 |
| Jewish General Hospital | Montréal | Québec | 67 |
| McMaster University | Hamilton | Ontario | 90 |
| Mount Sinai Hospital | Toronto | Ontario | 60 |
| Ottawa Hospital Research Institute | Ottawa | Ontario | 37 |
| St. Joseph's Health Care | London | Ontario | 81 |
| Toronto Western Hospital | Toronto | Ontario | 13 |
| University of Alberta | Edmonton | Alberta | 4 |
| University of British Columbia – Providence Health Care Research Institute | Vancouver | British Columbia | 37 |
| External Enrolment (Canada) | ·· | Canada | 9 |
| External Enrolment (English-speaking – not in North America) | ·· | | 2 |

| | | | |
|---|---|---|---|
| **France** | | | **590** |
| AP-HM, Hôpital Nord | Marseille | Provence-Alpes-Côte d'Azur | 38 |
| AP-HP Cochin | Paris | Ile de France | 290 |
| AP-HP St-Antoine | Paris | Ile de France | 36 |
| AP-HP St-Louis | Paris | Ile de France | 39 |
| CHU Lyon Sud | Lyon | Auvergne-Rhône-Alpes | 5 |
| CHU de Rouen | Rouen | Normandie | 74 |
| CHU de Dijon | Dijon | Bourgogne-Franche-Comté | 4 |
| CHU de Clermont Ferrand, Gabriel Montpied | Clermont-Ferrand | Auvergne-Rhône-Alpes | 4 |
| CHU de Lille Claude Huriez | Lille | Hauts de France | 40 |
| CHU de Nantes – Hôtel Dieu | Nantes | Pays de la Loire | 19 |
| CHU Strasbourg, Nouvel Hôpital Civil | Strasbourg | Grand Est | 26 |
| Uneos-Groupe Hospitalier Associatif Metz | Metz | Grand Est | 11 |
| External Enrolment (French-speaking – not in Canada) | .. | Europe | 4 |
| **Mexico** | | | **23** |
| Instituto Nacional de Ciencias Médicas y Nutrición Salvador Zubirán | Mexico City | Mexico City | 23 |
| **Spain** | | | **42** |
| Hospital 12 de Octubre Madrid | Madrid | Madrid | 42 |
| **UK** | | | **191** |
| Royal Free London NHS Foundation Trust | London | England | 132 |
| Salford Royal NHS Foundation Trust | Manchester | England | 59 |
| **USA** | | | **778** |

| | | | |
|---|---|---|---|
| Arthritis Associates of Southern California | Los Angeles | California | 67 |
| Columbia University | New York | New York | 1 |
| Georgetown University | Washington | Washington, DC | 63 |
| Hospital for Special Surgery | New York | New York | 30 |
| Johns Hopkins University | Baltimore | Maryland | 236 |
| Northwestern University | Chicago | Illinois | 52 |
| Stanford University | Palo Alto | California | 21 |
| University of California Los Angeles | Los Angeles | California | 52 |
| University of Michigan | Ann Arbor | Michigan | 127 |
| University of Pittsburgh | Pittsburgh | Pennsylvania | 23 |
| University of Texas – Houston | Houston | Texas | 41 |
| University of Utah | Salt Lake City | Utah | 55 |
| External Enrolment (USA) | ·· | USA | 10 |

AP-HM = Assistance Publique - Hôpitaux de Marseille; AP-HP = Assistance Publique - Hôpitaux de Paris; CHU = Centre Hospitalier Universitaire; NHS = National Health Service

**Appendix Figure 1.** Frequency distribution of the number of itch severity numerical rating scale assessments by past-week itch severity score.

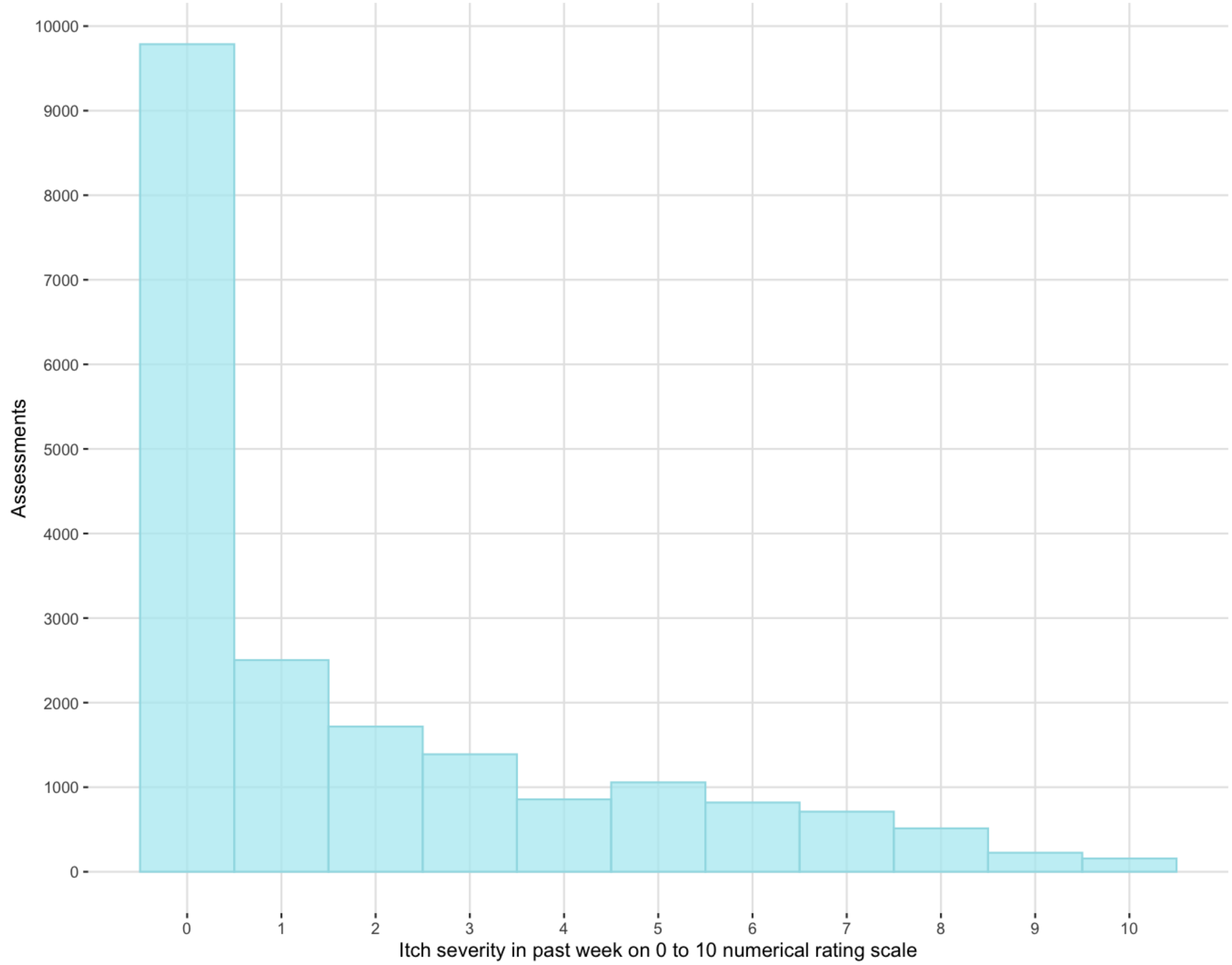

**Appendix Figure 2.** Predicted probability of itch (numerical rating scale score > 0) by age at time of assessment for different ages of non-Raynaud phenomenon symptom onset, with 95% confidence intervals.

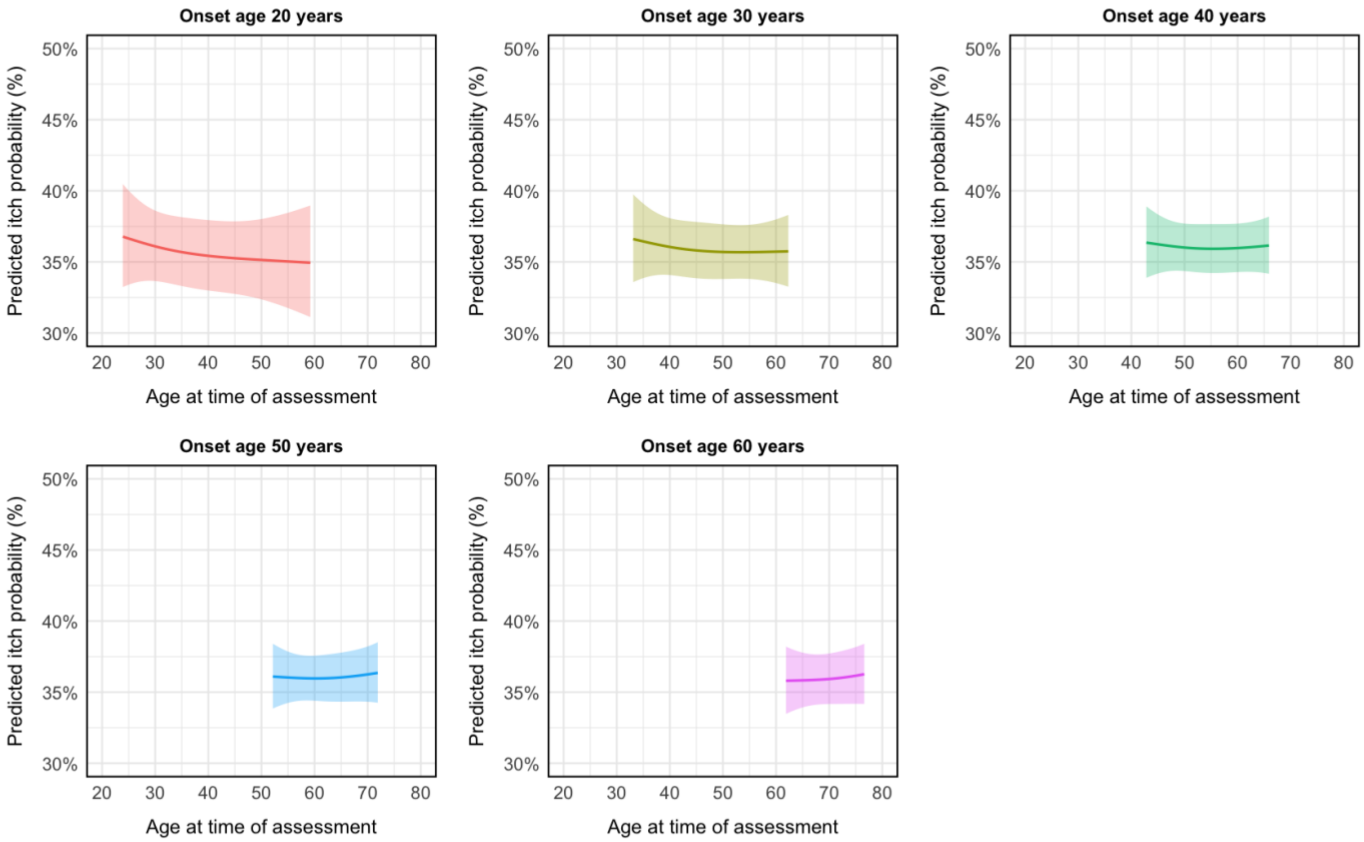


For each predicted trajectory (e.g., non-Raynaud phenomenon symptom onset age 30 years), we defined trajectory boundaries using participants with onset ages ±5 years from the prediction age (e.g., ≥25 to <35 years). We started each trajectory at the 10th percentile of time since non- Raynaud phenomenon symptom onset and finished at the 90th percentile to ensure adequate data for estimates. Resulting ranges of time since non-Raynaud phenomenon symptom onset in the figure are for onset age 20 years, 3·9 to 39·2 years; onset age 30 years, 3·1 to 32·3 years; onset age 40 years, 2·8 to 25·9 years; onset age 50 years, 2·2 to 21·9 years; and onset age 60 years, 1·9 to 16·6 years.

**Appendix Figure 3.** Predicted severity of past-week itch, if present, on 0 to 10 numerical rating scale by age at time of assessment for different ages of non-Raynaud phenomenon symptom onset, with 95% confidence intervals.

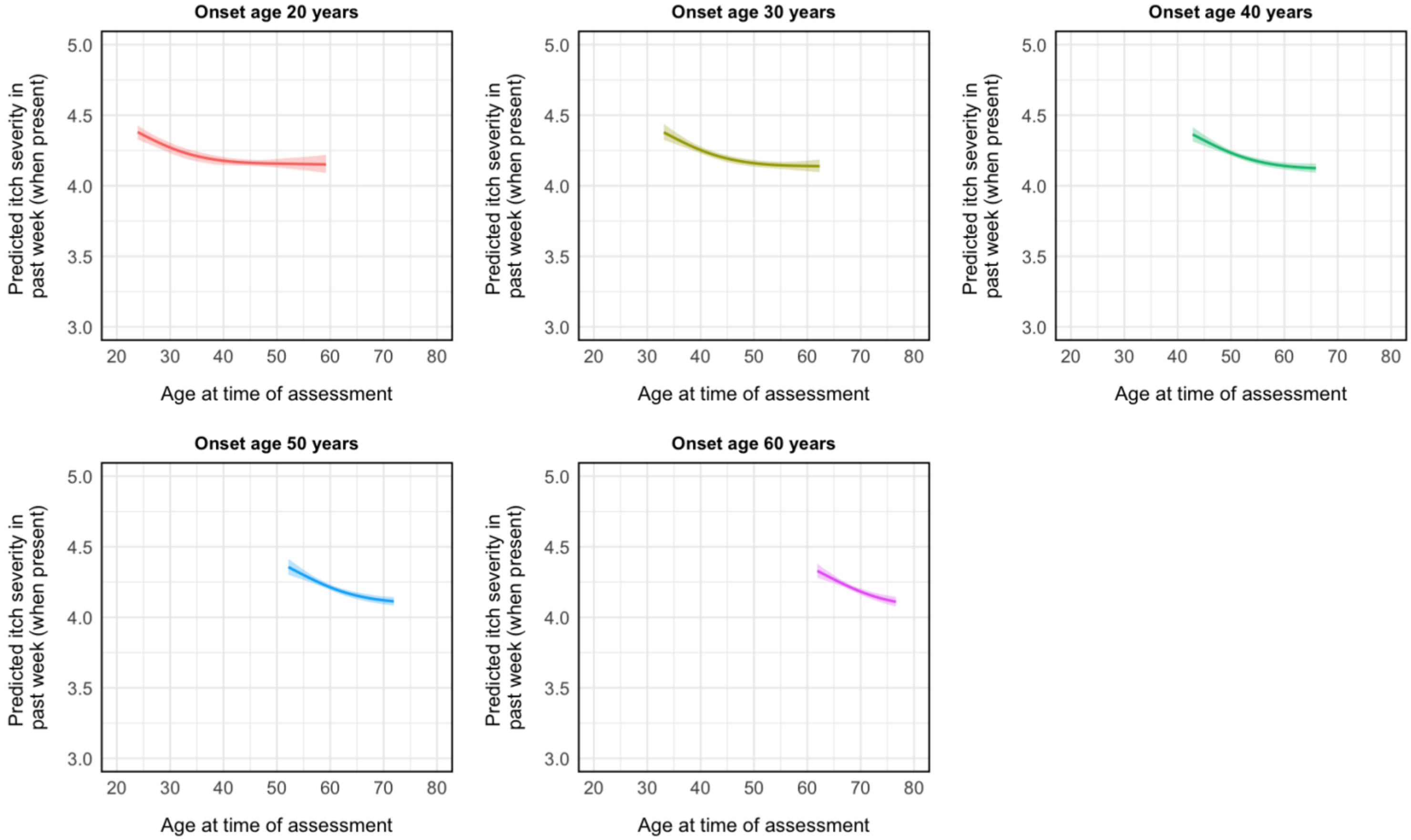


For each predicted trajectory (e.g., non-Raynaud phenomenon symptom onset age 30 years), we defined trajectory boundaries using participants with onset ages ±5 years from the prediction age (e.g., ≥25 to <35 years). We started each trajectory at the 10th percentile of time since non-Raynaud phenomenon symptom onset and finished at the 90th percentile to ensure adequate data for estimates. Resulting ranges of time since non-Raynaud phenomenon symptom onset in the figure are for onset age 20 years, 3·9 to 39·2 years; onset age 30 years, 3·1 to 32·3 years; onset age 40 years, 2·8 to 25·9 years; onset age 50 years, 2·2 to 21·9 years; and onset age 60 years, 1·9 to 16·6 years.

**Appendix Figure 4.** Predicted probability of itch (numerical rating scale score > 0) by age at time of assessment for different ages of non-Raynaud phenomenon symptom onset, including only participants who completed the itch numerical rating scale for the first time ≤ 10 years after non-Raynaud phenomenon symptom onset.

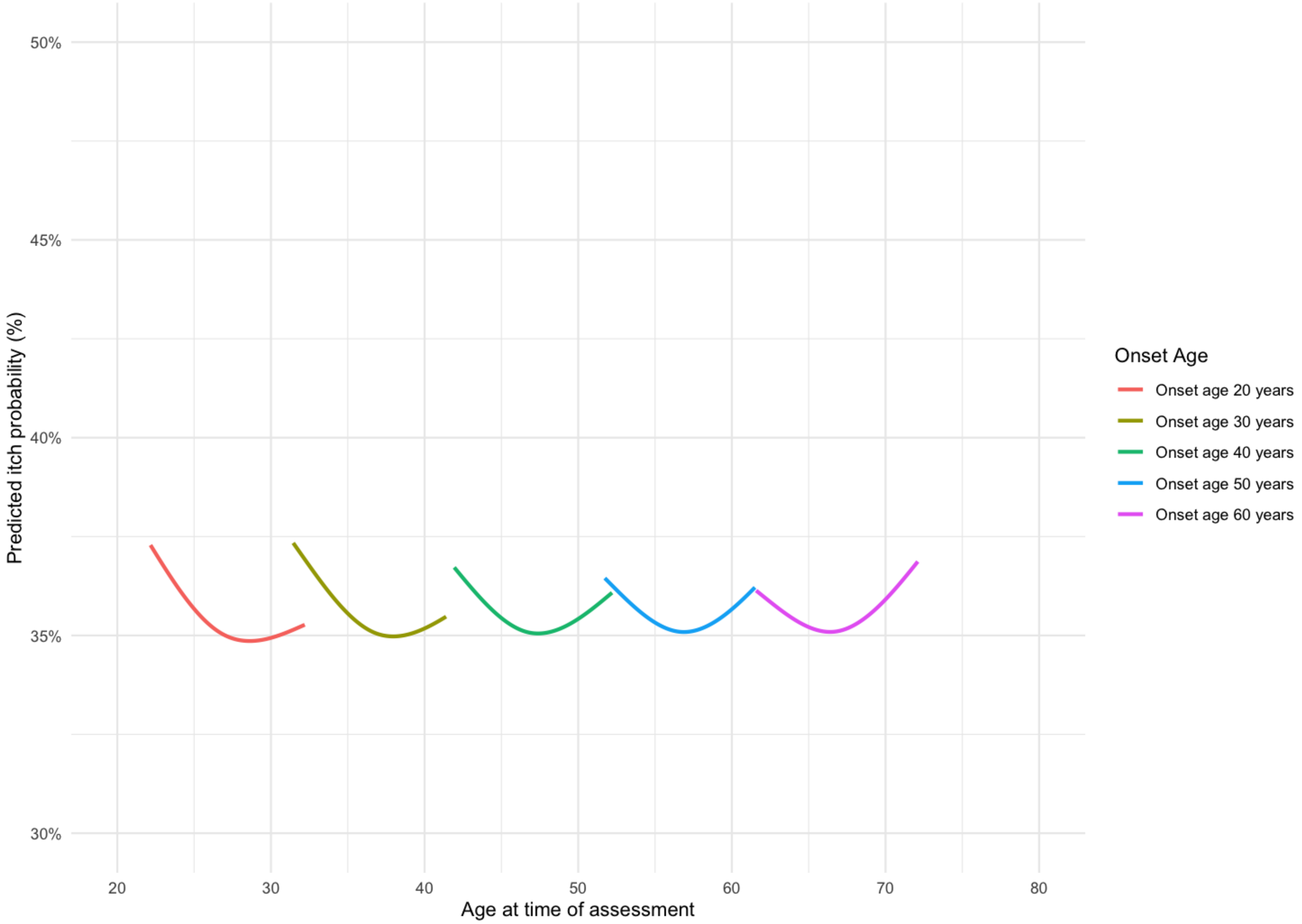


For each predicted trajectory (e.g., non-Raynaud phenomenon symptom onset age 30 years), we defined trajectory boundaries using participants with onset ages ± 5 years from the prediction age (e.g., 25 to 35 years). We started each trajectory at the 10$^{th}$ percentile of time since non-Raynaud phenomenon symptom onset and finished at the 90$^{th}$ percentile to ensure adequate data for estimates. Resulting ranges of time since non-Raynaud phenomenon symptom onset in the figure are for onset age for onset age 20 years, 2·2 to 12·2 years; onset age 30 years, 1·4 to 11·4 years; onset age 40 years, 1·9 to 12·2 years; onset age 50 years, 1·7 to 11·5 years; and onset age 60 years, 1·6 to 12·1 years.

**Appendix Figure 5.** Predicted severity of past-week itch, if present, on 0 to 10 numerical rating scale by age at time of assessment for different ages of non-Raynaud phenomenon symptom onset, including only participants who completed the itch numerical rating scale for the first time ≤ 10 years after non-Raynaud phenomenon symptom onset.

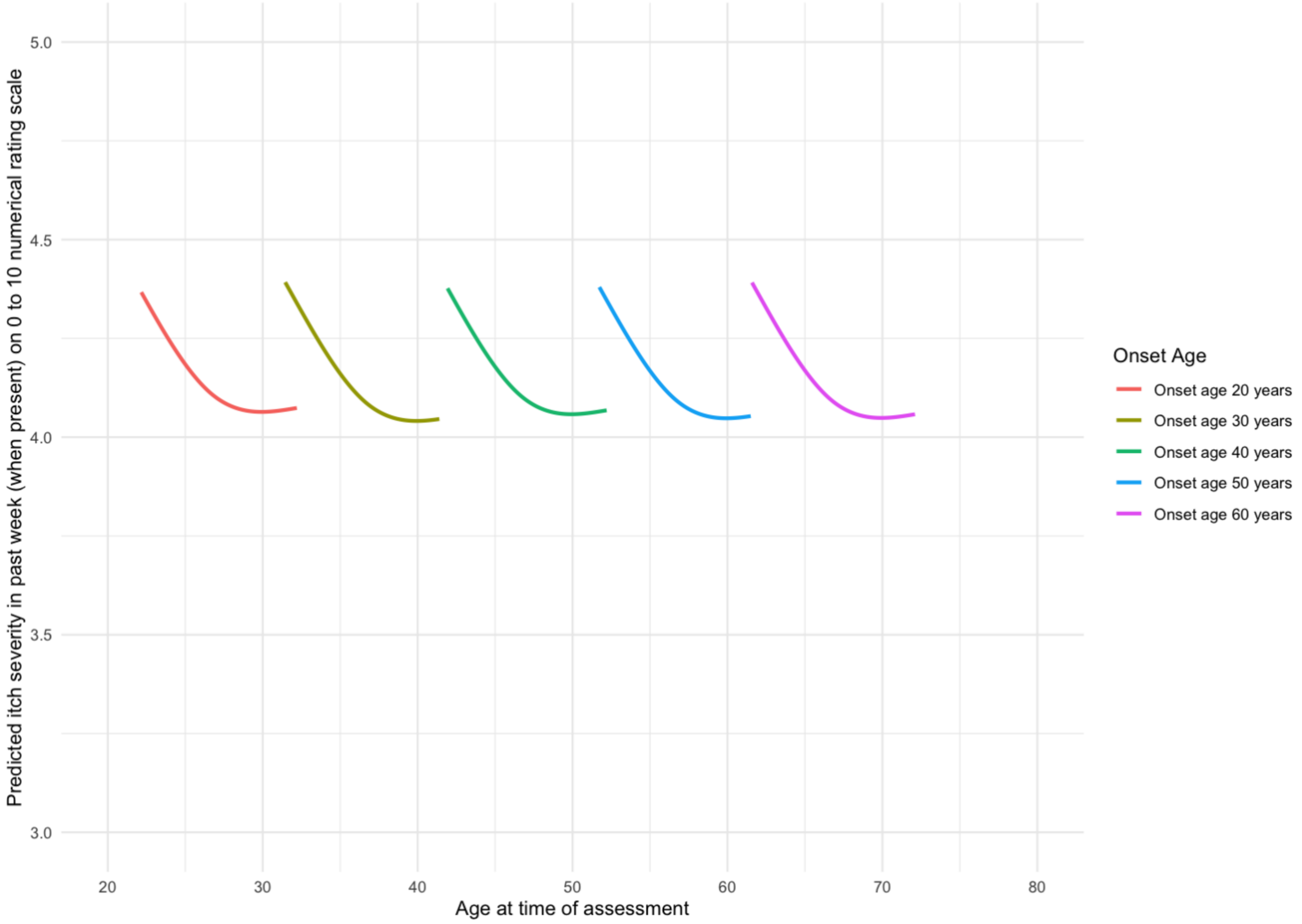


For each predicted trajectory (e.g., non-Raynaud phenomenon symptom onset age 30 years), we defined trajectory boundaries using participants with onset ages ± 5 years from the prediction age (e.g., 25 to 35 years). We started each trajectory at the 10$^{th}$ percentile of time since non- Raynaud phenomenon symptom onset and finished at the 90$^{th}$ percentile to ensure adequate data for estimates. Resulting ranges of time since non-Raynaud phenomenon symptom onset in the figure are for onset age for onset age 20 years, 2·2 to 12·2 years; onset age 30 years, 1·4 to 11·4 years; onset age 40 years, 1·9 to 12·2 years; onset age 50 years, 1·7 to 11·5 years; and onset age 60 years, 1·6 to 12·1 years.

**Appendix Table 2.** Predicted values of itch probability (numerical rating scale score > 0) and itch severity in past week (0 to 10 numerical rating scale), if present, for an average participant at 2 years, 3 years, 4 years, 5 years, 7 years and 10 years post non-Raynaud phenomenon symptom onset with 95% confidence intervals by age at non-Raynaud phenomenon symptom onset, including only participants who completed the itch numerical rating scale for the first time ≤ 10 years after non-Raynaud phenomenon symptom onset.

| | Age of non-Raynaud phenomenon symptom onset | | | | |
|---|---|---|---|---|---|
| | **20 years** | **30 years** | **40 years** | **50 years** | **60 years** |
| **Years since non-Raynaud phenomenon symptom onset[a]** | | | **Itch probability (%)** | | |
| 2 years | ·· | 37·0 (32·4 to 41·9) | 36·7 (33·0 to 40·5) | 36·3 (33·1 to 39·7) | 36·0 (32·6 to 39·5) |
| 3 years | 36·8 (31·9 to 41·9) | 36·5 (32·6 to 40·6) | 36·2 (33·1 to 39·4) | 36·0 (33·3 to 38·7) | 35·7 (32·9 to 38·6) |
| 4 years | 36·2 (32·0 to 40·6) | 36·0 (32·6 to 39·5) | 35·8 (33·1 to 38·6) | 35·6 (33·3 to 38·0) | 35·4 (33·0 to 37·9) |
| 5 years | 35·7 (31·9 to 39·6) | 35·6 (32·5 to 38·7) | 35·4 (32·9 to 38·0) | 35·3 (33·1 to 37·6) | 35·2 (32·9 to 37·6) |
| 7 years | 35·0 (31·5 to 38·7) | 35·0 (32·1 to 38·1) | 35·1 (32·6 to 37·6) | 35·1 (32·8 to 37·4) | 35·1 (32·8 to 37·5) |
| 10 years | 34·9 (31·0 to 39·1) | 35·2 (32·1 to 38·4) | 35·4 (33·0 to 37·9) | 35·7 (33·4 to 38·0) | 35·9 (33·3 to 38·6) |
| | **Itch severity in past week (when present) on 0 to 10 numerical rating scale** | | | | |
| 2 years | ·· | 4·4 (4·3 to 4·5) | 4·4 (4·3 to 4·5) | 4·4 (4·3 to 4·5) | 4·4 (4·3 to 4·5) |
| 3 years | 4·3 (4·2 to 4·4) | 4·3 (4·2 to 4·4) | 4·3 (4·2 to 4·4) | 4·3 (4·2 to 4·4) | 4·3 (4·2 to 4·4) |
| 4 years | 4·2 (4·2 to 4·3) | 4·2 (4·2 to 4·3) | 4·2 (4·2 to 4·3) | 4·2 (4·2 to 4·3) | 4·2 (4·2 to 4·3) |
| 5 years | 4·2 (4·1 to 4·2) | 4·2 (4·1 to 4·2) | 4·2 (4·1 to 4·2) | 4·2 (4·1 to 4·2) | 4·2 (4·1 to 4·2) |
| 7 years | 4·1 (4·1 to 4·1) | 4·1 (4·0 to 4·1) | 4·1 (4·0 to 4·1) | 4·1 (4·0 to 4·1) | 4·1 (4·0 to 4·1) |
| 10 years | 4·1 (4·0 to 4·1) | 4·0 (4·0 to 4·1) | 4·1 (4·0 to 4·1) | 4·0 (4·0 to 4·1) | 4·0 (4·0 to 4·1) |

[a]For each non-Raynaud phenomenon symptom onset age (e.g., 30 years), we defined boundaries using participants with onset ages ±5 years from the prediction age (e.g., ≥25 to <35 years). We reported predicted values from the 10th percentile of years since non-Raynaud phenomenon symptom onset at the first assessment to the 90th percentile at the last assessment to ensure adequate data for estimates.

**Appendix Table 3.** Predictors of itch presence (numerical rating scale score > 0) with 95% confidence intervals from main analysis (N = 2173 participants, 19733 assessments) and sensitivity analysis including only participants who completed the itch numerical rating scale for the first time ≤ 10 years after non-Raynaud phenomenon symptom onset (N = 1192 participants, 10549 assessments).

| **Variable** | **Odds ratio for itch presence** | |
|---|---|---|
| | **Main analysis<br>N = 2173<br>Estimate (95% CI)** | **Sensitivity analysis<br>N = 1192<br>Estimate (95% CI)** |
| Basis spline (time since non-Raynaud phenomenon symptom onset, per 5 years) #1[a] | 0·46 (0·28 to 0·76) | 0·35 (0·17 to 0·70) |
| Basis spline (time since non-Raynaud phenomenon symptom onset, per 5 years) #2[a] | 0·61 (0·46 to 0·82) | 0·71 (0·45 to 1·11) |
| Age of non-Raynaud phenomenon symptom onset, per 10 years | 0·94 (0·89 to 0·99) | 0·92 (0·84 to 1·00) |
| Interaction: age of non-Raynaud phenomenon symptom onset x time since onset | 1·03 (1·01 to 1·05) | 1·07 (1·01 to 1·13) |
| Male sex (reference = female) | 1·06 (0·95 to 1·18) | 0·97 (0·84 to 1·10) |
| Country (reference = USA) | | |
| Canada | 1·45 (1·33 to 1·58) | 1·59 (1·41 to 1·78) |
| UK | 1·78 (1·57 to 2·02) | 1·33 (1·11 to 1·58) |
| France | 1·34 (1·22 to 1·47) | 1·34 (1·18 to 1·51) |
| Other (Australia, Mexico, Spain) | 1·59 (1·32 to 1·92) | 1·05 (0·80 to 1·38) |
| Diffuse cutaneous disease subtype (reference = limited cutaneous or sine scleroderma) | 1·25 (1·16 to 1·34) | 1·57 (1·43 to 1·73) |

CI = confidence interval
[a]Cubic spline parameter for years since non-Raynaud phenomenon symptom onset

**Appendix Table 4.** Predictors of past-week itch severity, if present, on 0 to 10 numerical rating scale with 95% confidence intervals from main analysis (N = 2173 participants, 19733 assessments) and sensitivity analysis including only participants who completed the itch numerical rating scale for the first time ≤ 10 years after non-Raynaud phenomenon symptom onset (N = 1192 participants, 10549 assessments).

| **Variable** | **Percent Difference in severity if present[a]** | |
|---|---|---|
| | **Main analysis<br>N = 2173<br>Estimate (95% CI)** | **Sensitivity analysis<br>N = 1192<br>Estimate (95% CI)** |
| Basis spline (time since non-Raynaud phenomenon symptom onset, per 5 years) #1[b] | -8·70 (-13·89 to -3·20) | -11·73 (-18·09 to -4·87) |
| Basis spline (time since non-Raynaud phenomenon symptom onset, per 5 years) #2[b] | -3·63 (-6·80 to -0·36) | -3·56 (-7·57 to 0·62) |
| Age of non-Raynaud phenomenon symptom onset, per 10 years | -7·07 (-11·42 to -2·50) | -8·40 (-13·65 to -2·84) |
| Male sex (reference = female) | -0·79 (-3·92 to 2·43) | -1·47 (-5·36 to 2·59) |
| Country (reference = USA) | | |
| Canada | -1·84 (-4·37 to 0·77) | -1·88 (-5·22 to 1·58) |
| UK | 0·80 (-2·75 to 4·48) | 2·23 (-2·99 to 7·74) |
| France | 3·28 (0·33 to 6·32) | 3·77 (-0·13 to 7·82) |
| Other (Australia, Mexico, Spain) | 2·07 (-3·59 to 8·05) | 0·55 (-8·16 to 10·09) |
| Diffuse cutaneous disease subtype (reference = limited cutaneous or sine scleroderma) | -0·69 (-2·87 to 1·53) | -0·53 (-3·40 to 2·42) |

CI = confidence interval

[a]Difference is multiplicative (e.g., 1.00 = 1% change). Positive numbers reflect greater itch severity.

[b]Cubic spline parameter for years since non-Raynaud phenomenon symptom onset

**Appendix Figure 6.** Predicted probability of itch (numerical rating scale score > 0) by age at time of assessment for different ages of non-Raynaud phenomenon symptom onset, by sex.

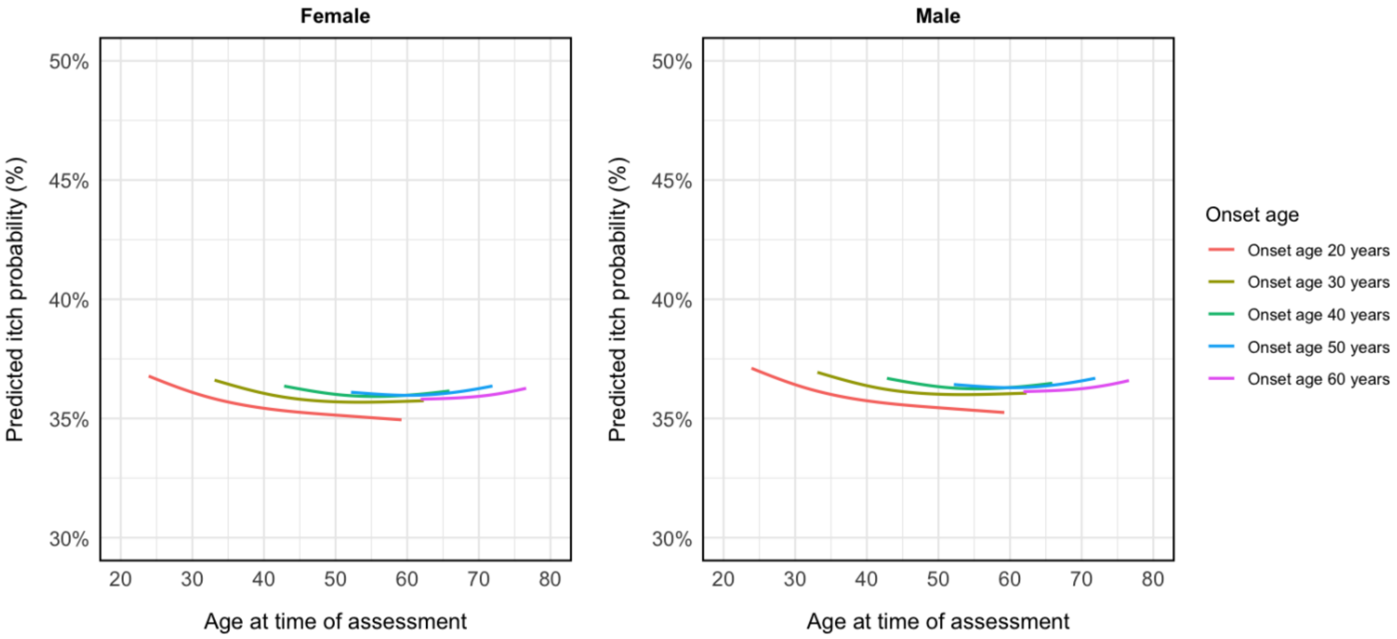

**Appendix Figure 7.** Predicted severity of past-week itch, if present, on 0 to 10 numerical rating scale by age at time of assessment for different ages of non-Raynaud phenomenon symptom onset, by sex.

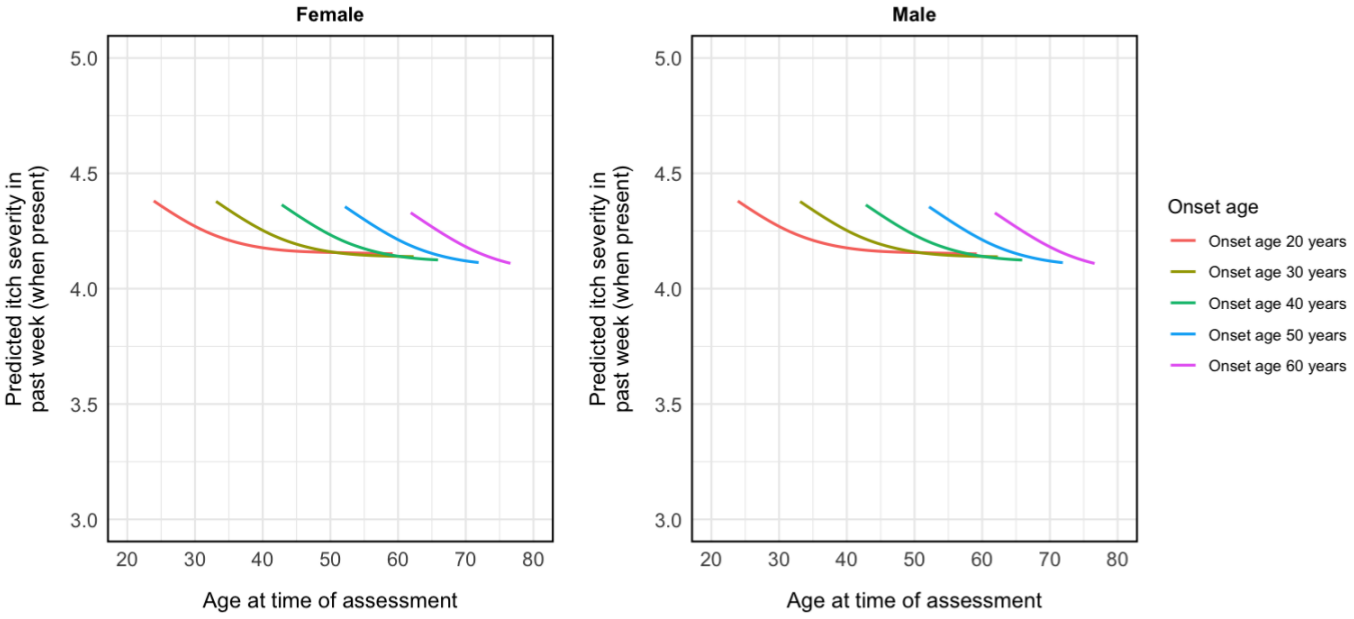

**Appendix Figure 8.** Predicted probability of itch (numerical rating scale score > 0) by age at time of assessment for different ages of non-Raynaud phenomenon symptom onset, by country.

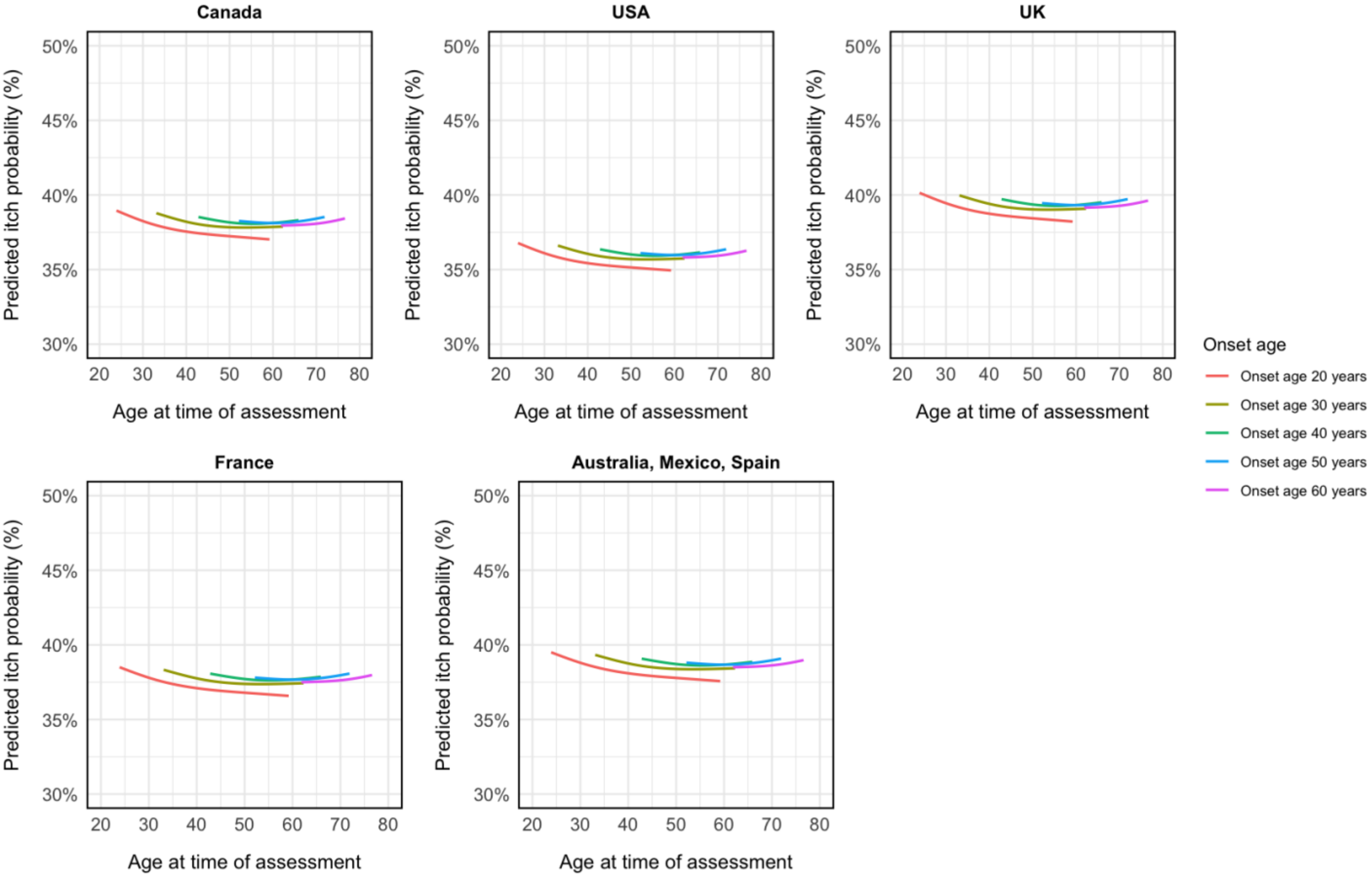

**Appendix Figure 9.** Predicted severity of past-week itch, if present, on 0 to 10 numerical rating scale by age at time of assessment for different ages of non-Raynaud phenomenon symptom onset, by country.

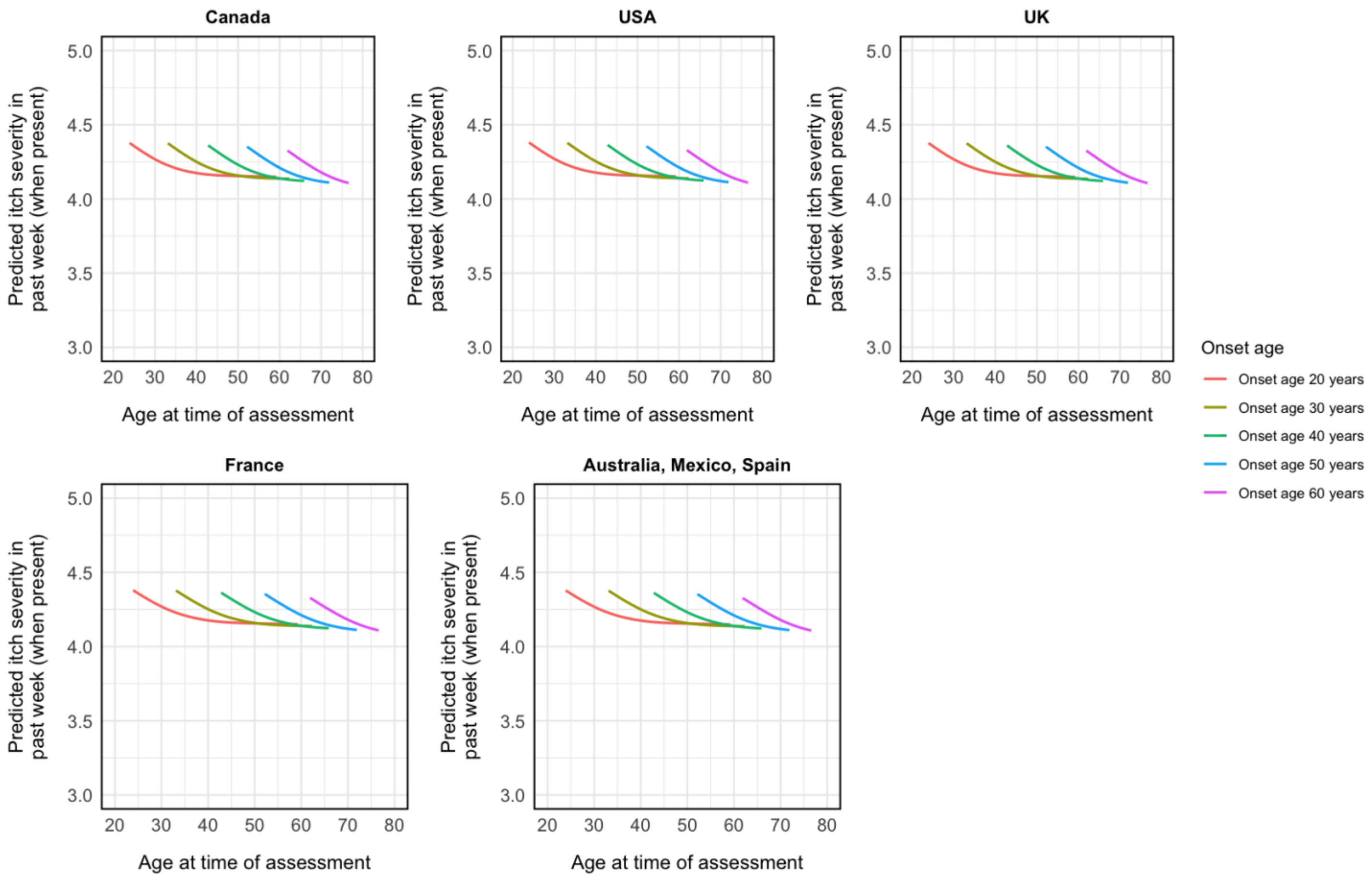

**Appendix Figure 10.** Predicted probability of itch (numerical rating scale score > 0) by age at time of assessment for different ages of non-Raynaud phenomenon symptom onset, by disease subtype.

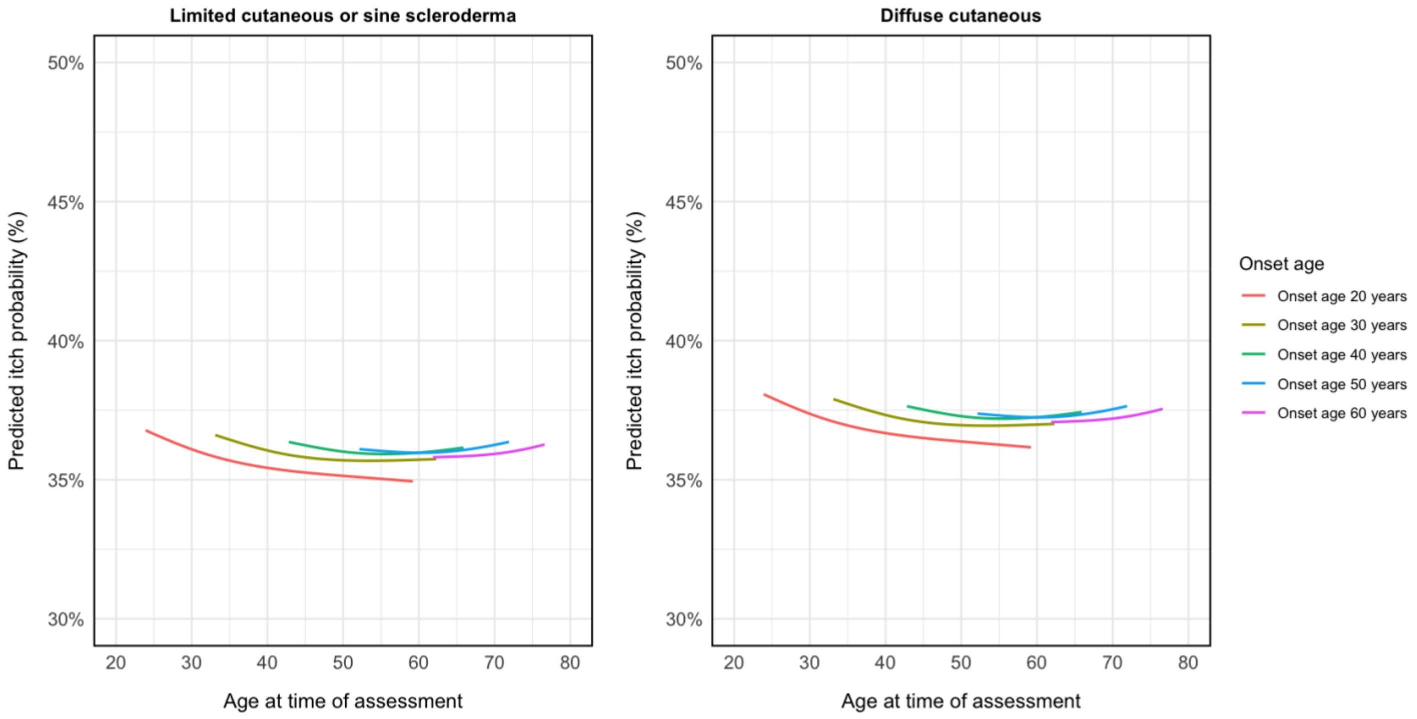

**Appendix Figure 11.** Predicted severity of past-week itch, if present, on 0 to 10 numerical rating scale by age at time of assessment for different ages of non-Raynaud phenomenon symptom onset, by disease subtype.

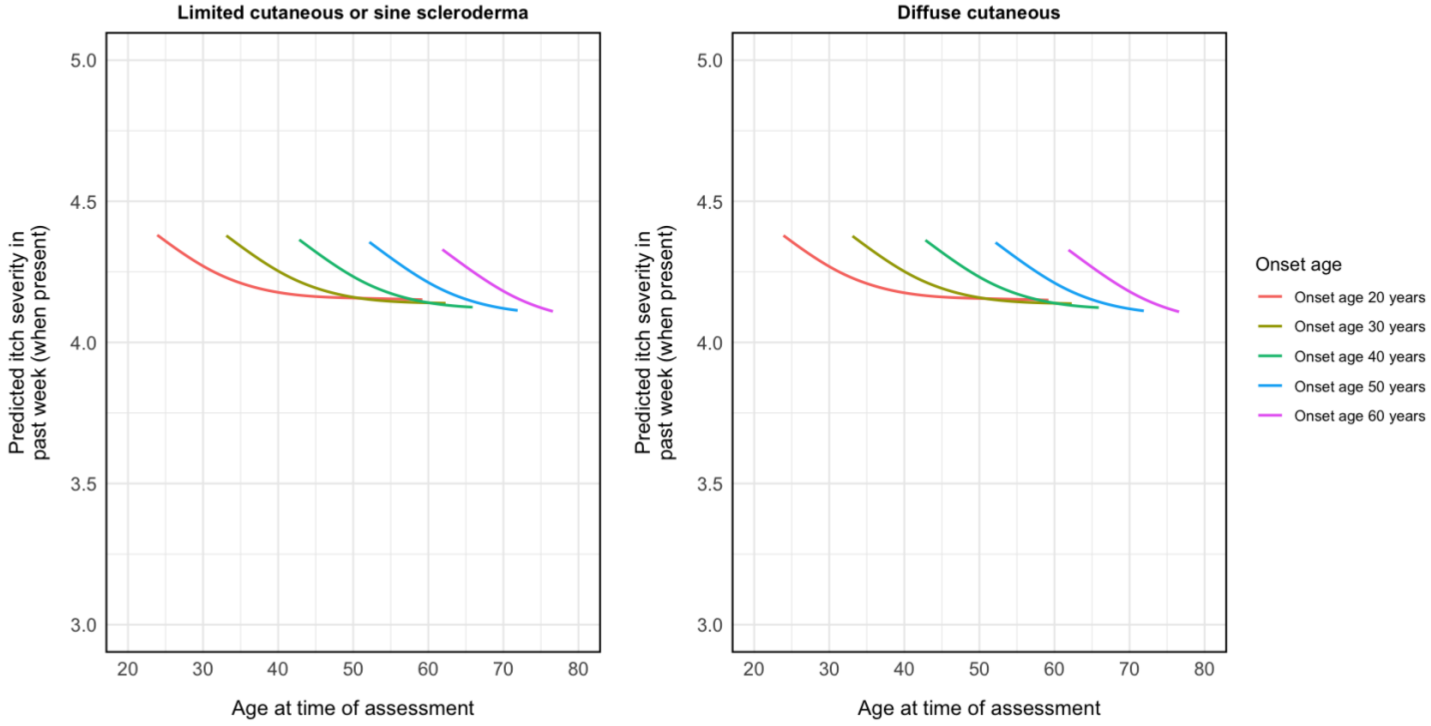
